# Resilience Beyond Diagnosis: Prospective Neural Correlates of Better-Than-Expected Outcomes in Depression

**DOI:** 10.64898/2025.12.28.25342267

**Authors:** Vincent Hammes, Katharina Brosch, Paula Usemann, Friederike David, Hannah Scherer, Christina Schmitter, Jochen Bauer, Tiana Borgers, Kira Flinkenflügel, Dominik Grotegerd, Elisabeth Leehr, Susanne Meinert, Frederike Stein, Lea Teutenberg, Florian Thomas-Odenthal, Udo Dannlowski, Tim Hahn, Hamidreza Jamalabadi, Andreas Jansen, Robert Miller, Igor Nenadić, Benjamin Straube, Albrecht Stroh, Tilo Kircher, Nina Alexander

## Abstract

**Background:** Resilience, the ability to adapt positively in the face of adversity, is shaped by the interplay of risk and protective factors. Previous magnetic resonance imaging (MRI) studies on resilience have predominantly focused on single factors, often operationalizing resilience dichotomously as the absence of psychiatric disorders despite adversity.

**Methods:** In this prospective MRI study, we defined resilience as “better-than-expected” depressive symptom severity (Hamilton Depression Rating Scale) relative to cumulative risk. Using ridge-regularized regression in N=1,804 participants (955 healthy, 849 depressed) from the Marburg-Münster Affective Disorders Cohort Study, we predicted symptom severity and derived residuals as measures of resilience. Residuals were then used to predict gray matter volume (GMV) and cortical thickness at baseline (T1) and two-year follow-up (T2; N=808), using voxel- and surface-based analyses. This approach was complemented by extreme-group comparisons of resilient (better-than-expected outcome) and vulnerable (worse-than-expected outcome) individuals.

**Results:** Cumulative risk explained 51.4% of variance in depressive symptoms at T1 and 44.2% at T2. Residual scores showed moderate stability over time (*r*=0.31, *p*<0.001). Region-of-interest and whole-brain analyses revealed no morphometric associations with resilience at T1. In contrast, higher resilience at T1 predicted lower GMV in the left inferior orbitofrontal gyrus (IOFG) and temporal pole at T2 (ROI, *p*_FWE(peak)_=0.002, partial *r*=0.18), with no changes in cortical thickness.

**Conclusion:** Resilience to cumulative risk, defined as better-than-expected depressive symptom severity, was not associated with immediate brain structural differences. However, prospective analyses revealed smaller GMV in the IOFG and temporal pole over time, potentially reflecting greater neural efficiency or delayed biological costs.

## Introduction

Resilience refers to the ability to adapt positively to stress and adversity, enabling individuals to maintain or regain mental health despite risk exposure. Rather than a fixed trait, resilience is considered a dynamic process shaped by the interaction of biological and environmental factors (1–3). Similarly, contemporary models of mental disorders, including the diathesis-stress framework, emphasize how cumulative exposures shape mental health outcomes such as Major Depressive Disorder (MDD) (4–7). Within this framework, both vulnerability and resilience to depression emerge from the cumulative and reciprocal effects of several, multi-systemic risk and protective factors over time (1,3,8,9).

Neural correlates of resilience to psychiatric disorders, particularly MDD and Post-traumatic Stress Disorder (PTSD), have been extensively studied. Across methodologically diverse studies, structural and functional alterations in prefrontal and cortico-limbic regions involved in stress and emotion regulation are repeatedly implicated (10–12). Most consistently, the medial prefrontal cortex (mPFC), encompassing the medial frontal gyrus, the anterior cingulate cortex (ACC) and the subcallosal area, as well as the orbitofrontal cortex (OFC) and hippocampus have been associated with resilience-related differences in brain structure (11,13–17). Less consistent findings involve additional prefrontal and limbic areas, such as the middle frontal gyrus, the superior frontal gyrus, and the amygdala (13,14,16,18,19). The majority of studies report larger or preserved gray matter volume (GMV) in these regions among resilient individuals, supporting the notion that resilience involves the maintenance or increase of structural integrity. This view is further substantiated by findings from the clinical literature, which link both MDD and PTSD to reduced GMV and cortical thickness in overlapping brain regions (20–23). However, some studies have reported null or even opposite associations, with smaller GMV observed in resilient individuals (16). This heterogeneity likely reflects differences in participant characteristics, resilience definitions, risk exposure, comparison groups, and sample size (11,15–17).

Alongside inconsistent findings, neuroimaging studies on resilience face several recurring conceptual and methodological challenges. First, most prior studies rely on cross-sectional designs, which limit the ability to draw conclusions about temporal dynamics or delayed neural effects. Second, resilience is often studied as the positive adaptation to single risk factors. However, the psychological impact of any given stressor depends on the broader accumulation of risk and protective factors that can either exacerbate or buffer its effect (7,24). Without accounting for cumulative risk profiles, an ostensibly “resilient” outcome may in fact reflect the absence of additional, unmeasured stressors – rendering these individuals effectively low-risk rather than genuinely resilient (25). Indeed, several studies have demonstrated that focusing on single risk factors fails to capture the complexity of resilient adaptation, emphasizing the need for comprehensive, multimodal assessments that integrate multiple resilience-related influences (26–28). In line with this, risk factors can exert both distinct and interacting effects on brain morphometry in a dose-dependent manner, whereas protective factors may buffer or reverse stress-related alterations (29–33).

Third, the commonly used dichotomous, diagnosis-based operationalization – defining resilience as the absence of a psychiatric disorder despite adversity – has two key drawbacks. In particular, comparisons between healthy (resilient) and depressed (non-resilient) individuals cannot disentangle disorder-driven brain alterations from those potentially linked to resilience. In such designs, larger GMV or cortical thickness in the resilient group may simply reflect disorder-associated reductions in the clinical group, rather than resilience-specific features. In fact, studies including healthy, low-risk comparison groups have shown that larger GMV in resilient individuals is evident only relative to the diagnosed group, and not compared to low-risk controls, where no differences or even reductions in GMV have emerged (16,34). Moreover, risk exposure, independent of clinical diagnosis, can leave measurable traces on brain structure (15,29,31,35,36). As such, individuals deemed resilient may nonetheless exhibit neural alterations shaped by their risk history, underscoring the importance of accounting for cumulative adversity in resilience research. In addition, a resilient outcome is not simply the absence of a diagnosis. Adaptive functioning under adversity depends on the level of cumulative risk at the time of stressor exposure. A dichotomous classification cannot account for variation in symptom severity and may overlook individuals who, despite developing depressive symptoms, show better outcomes than expected given their risk load (24,25,37,38).

In sum, resilience-focused neuroimaging research has largely employed diagnosis-based comparisons, often without accounting for individuals’ cumulative risk profiles or incorporating prospective assessments. To address these constraints, we built on an established residual-based framework that conceptualizes resilience as a *better-than-expected* symptom outcome given multi-systemic risk exposure (24,25,39–41). Within this framework, depressive symptom severity is modeled as a function of environmental and psychosocial risk and protective factors, and individual deviations from predicted outcomes are interpreted as continuous indices of resilience beyond diagnostic status. We applied this framework to a large, deeply phenotyped cohort of patients with MDD and healthy controls, with structural magnetic resonance imaging (MRI) assessed at baseline and two-year follow-up, enabling a dimensional and prospective investigation of neural correlates of resilience.

## Methods and Materials

### Sample

Participants were drawn from the Marburg–Münster Affective Disorders Cohort Study (MACS) within the FOR2107 consortium (42). From this cohort, we included N=1,804 adults with available MRI data at baseline (955 HC, 849 MDD). A subset (N=808; 442 HC, 366 MDD) completed follow-up MRI assessment approximately two years after baseline (T1→T2; mean interval=2.2 years, SD=0.3). Participants were 18–65 years old (M=35.2 years, SD=13.2), 65% female and of Central-European ancestry.

Recruitment was conducted via newspapers, advertisements and flyers. Patients were additionally recruited from in- and out-patient departments of the Universities of Marburg and Münster and affiliated psychiatric hospitals. Participants underwent deep phenotyping using a comprehensive questionnaire battery, neuropsychological assessment, biomaterial collection, and functional and structural neuroimaging (see 42). The German version of the structured clinical interview for the DSM-IV-TR (SCID-I) (43) was administered on site by trained staff. Exclusion criteria were current substance dependence, lifetime neurological or severe somatic disorders, traumatic head injury, verbal intelligence quotient (IQ) < 80, and standard MRI contraindications (e.g., pregnancy, ferromagnetic implants). HC met no criteria for any current or lifetime DSM-IV-TR psychiatric disorder; MDD participants met criteria for a current or lifetime diagnosis of MDD. Participants provided written informed consent and received financial compensation. The study adhered to the Declaration of Helsinki and was approved by the local ethics committees of Marburg and Münster, Germany.

### Assessment of Risk, Protective, and Outcome Variables

Depressive symptom severity was modeled dimensionally using the interviewer-rated 17-item Hamilton Depression Rating Scale (HAM-D). Items are rated on 3– or 5-point scales and summed to a total score (range 0–52), with higher scores indicating greater symptom severity (44).

As predictors of depressive symptom severity, we included 22 well-established risk and protective factors routinely assessed in the MACS cohort, covering familial psychiatric risk, childhood adversity, recent stressful life events (positive/negative; impact-weighted), perceived stress, social resources (perceived support and frequency of social interactions), personality (Big Five), adult attachment, cognitive ability (verbal/vocabulary-based IQ), socioeconomic status (education, household income) and immigration. Table 1 lists all constructs and instruments. The theoretical and empirical rationale for factor selection, together with full construct definitions and scoring details is provided in the Supplement 1.

**Table 1.**
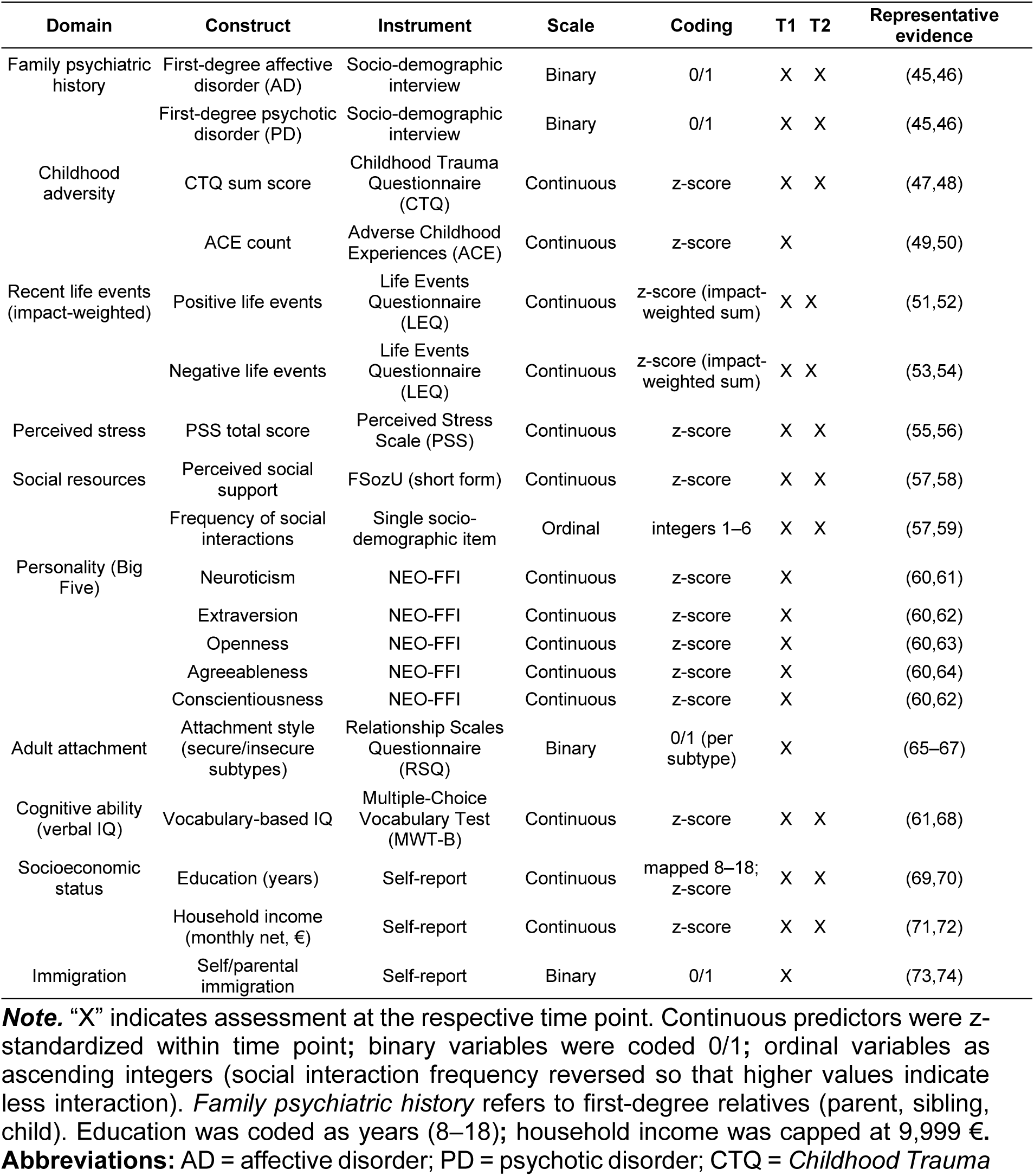
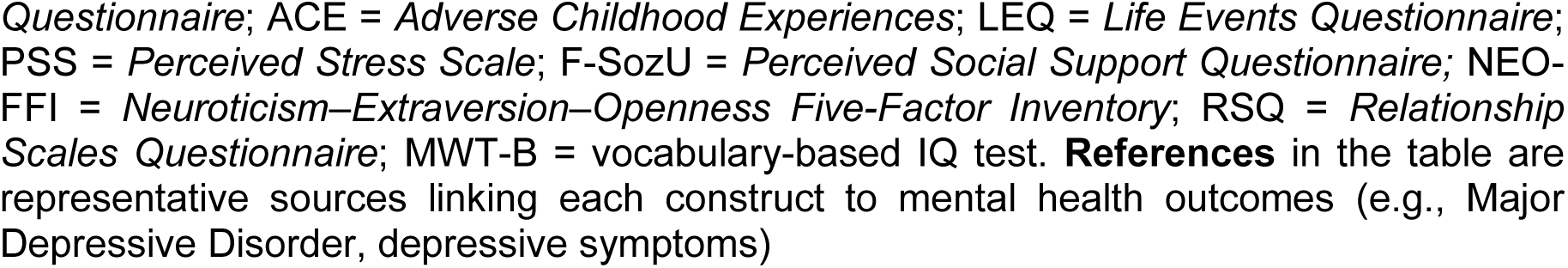
List of Risk and Protective Factors Included at T1 and T2.

| Domain | Construct | Instrument | Scale | Coding | T1 | T2 | Representative evidence |
| --- | --- | --- | --- | --- | --- | --- | --- |
| Family psychiatric history | First-degree affective disorder (AD) | Socio-demographic interview | Binary | 0/1 | X | X | (45,46) |
|  | First-degree psychotic disorder (PD) | Socio-demographic interview | Binary | 0/1 | X | X | (45,46) |
| Childhood adversity | CTQ sum score | Childhood Trauma Questionnaire (CTQ) | Continuous | z-score | X | X | (47,48) |
|  | ACE count | Adverse Childhood Experiences (ACE) | Continuous | z-score | X |  | (49,50) |
| Recent life events (impact-weighted) | Positive life events | Life Events Questionnaire (LEQ) | Continuous | z-score (impact-weighted sum) | X | X | (51,52) |
|  | Negative life events | Life Events Questionnaire (LEQ) | Continuous | z-score (impact-weighted sum) | X | X | (53,54) |
| Perceived stress | PSS total score | Perceived Stress Scale (PSS) | Continuous | z-score | X | X | (55,56) |
| Social resources | Perceived social support | FSozU (short form) | Continuous | z-score | X | X | (57,58) |
|  | Frequency of social interactions | Single socio-demographic item | Ordinal | integers 1–6 | X | X | (57,59) |
| Personality (Big Five) | Neuroticism | NEO-FFI | Continuous | z-score | X |  | (60,61) |
|  | Extraversion | NEO-FFI | Continuous | z-score | X |  | (60,62) |
|  | Openness | NEO-FFI | Continuous | z-score | X |  | (60,63) |
|  | Agreeableness | NEO-FFI | Continuous | z-score | X |  | (60,64) |
|  | Conscientiousness | NEO-FFI | Continuous | z-score | X |  | (60,62) |
| Adult attachment | Attachment style (secure/insecure subtypes) | Relationship Scales Questionnaire (RSQ) | Binary | 0/1 (per subtype) | X |  | (65–67) |
| Cognitive ability (verbal IQ) | Vocabulary-based IQ | Multiple-Choice Vocabulary Test (MWT-B) | Continuous | z-score | X | X | (61,68) |
| Socioeconomic status | Education (years) | Self-report | Continuous | mapped 8–18; z-score | X | X | (69,70) |
|  | Household income (monthly net, €) | Self-report | Continuous | z-score | X | X | (71,72) |
| Immigration | Self/parental immigration | Self-report | Binary | 0/1 | X |  | (73,74) |

**Note.** “X” indicates assessment at the respective time point. Continuous predictors were z-standardized within time point; binary variables were coded 0/1; ordinal variables as ascending integers (social interaction frequency reversed so that higher values indicate less interaction). *Family psychiatric history* refers to first-degree relatives (parent, sibling, child). Education was coded as years (8–18); household income was capped at 9,999 €. **Abbreviations:** AD = affective disorder; PD = psychotic disorder; CTQ = *Childhood Trauma*
Questionnaire; ACE = Adverse Childhood Experiences; LEQ = Life Events Questionnaire; PSS = Perceived Stress Scale; F-SozU = Perceived Social Support Questionnaire; NEO-FFI = Neuroticism–Extraversion–Openness Five-Factor Inventory; RSQ = Relationship Scales Questionnaire; MWT-B = vocabulary-based IQ test. **References** in the table are representative sources linking each construct to mental health outcomes (e.g., Major Depressive Disorder, depressive symptoms)

### Operationalization of Residual-Based Resilience

By applying a residual approach, this study employs a data-driven operationalization of resilience as a “better-than-expected” level of depressive symptoms given an individual’s cumulative risk. We included the multi-system risk and protective factors described above as predictors of depressive symptom severity (HAM-D sum score), with sex and age included as covariates. At baseline (T1), we fit a ridge-regularized linear regression model (*glmnet,* v4.1-8; 74) while z-standardizing continuous predictors and categorical variables entered as dummy-coded factors (Table 1) to estimate the expected HAM-D score (“expected symptom severity”) for each individual (Figure 1). In a secondary stability analysis, residuals were re-estimated at T2 by substituting updated T2 measures where available (Table 1) and carrying forward T1 values for stable constructs not reassessed at T2 (Figure S1). Ridge-regularized regression was chosen to retain all predictors while controlling for multicollinearity and ensuring non-zero shrinkage of included variables. Model tuning used 10-fold cross-validation, with each fold serving once as the validation set and nine times as part of the training set. The optimal regularization parameter λ was determined as λ_min_ (i.e., the smallest mean cross-validated error, MSE). To enhance generalizability, the more parsimonious λ_SE_ — the largest λ within one standard error of λ_min_ — was selected for the final model. Predicted HAM-D scores were calculated as the sum of the products of each predictor and its regularized regression coefficient plus the intercept. Residual-based resilience scores were computed as the difference between observed and predicted HAM-D score and thus span a continuous dimension from resilience (negative residuals; better-than-expected) to vulnerability (positive residuals; worse-than-expected).

**Figure 1.**
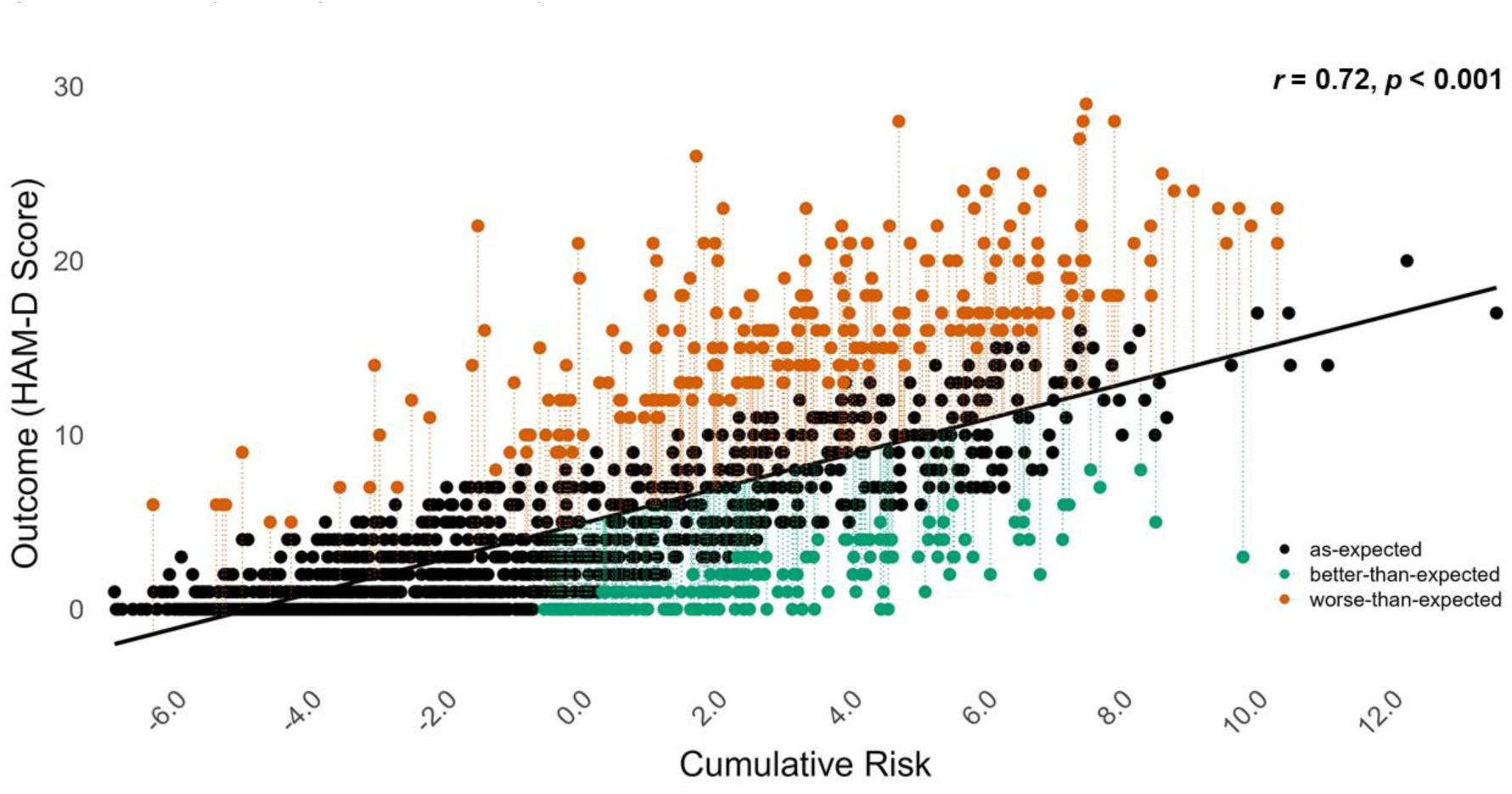
Ridge-Regularized Regression Model at T1. ***Note.*** Ridge regression at T1 showing the significant positive association between cumulative risk and depressive symptom severity (HAM-D, *r* = 0.72, *p* < 0.001). Residuals reflect individual deviations from predicted symptom scores and index resilience (green; better-than-expected) and vulnerability (red; worse-than-expected), defined as values exceeding ±1 residual standard error (RSE = 4.18).

Implementation of the statistical framework and all non-imaging analyses were performed in *R* (v4.4.2). Parameter settings and preprocessing steps are detailed in the openly available analysis code (76).

### Extreme Group Classification

Extreme groups were operationalized from the T1 residual distribution using a symmetric ±1 residual standard error (RSE=4.18; Figure 1). Participants with residuals < −4.18 (observed–expected HAM-D; better-than-expected) formed the resilient group, and those with residuals > 4.18 (worse-than-expected) the vulnerable group. This yielded n=208 resilient and n=251 vulnerable participants at T1 (mean residuals=−5.70 and 8.02, respectively); n=95 and n=97 were reassessed at T2.

### Clinical and Demographic Group Comparisons

Clinical and demographic characteristics were compared between the resilient and vulnerable extreme groups (Table 2). Continuous variables were analyzed with two-sample t-tests when assumptions of normality and homogeneity of variances were met. Otherwise, Wilcoxon rank-sum tests with Bonferroni correction for multiple comparisons were used. Categorical variables were tested with Pearson’s χ² tests or Fisher’s exact tests when expected cell counts were small. All tests were two-sided with *α*=0.05.

**Table 2.**
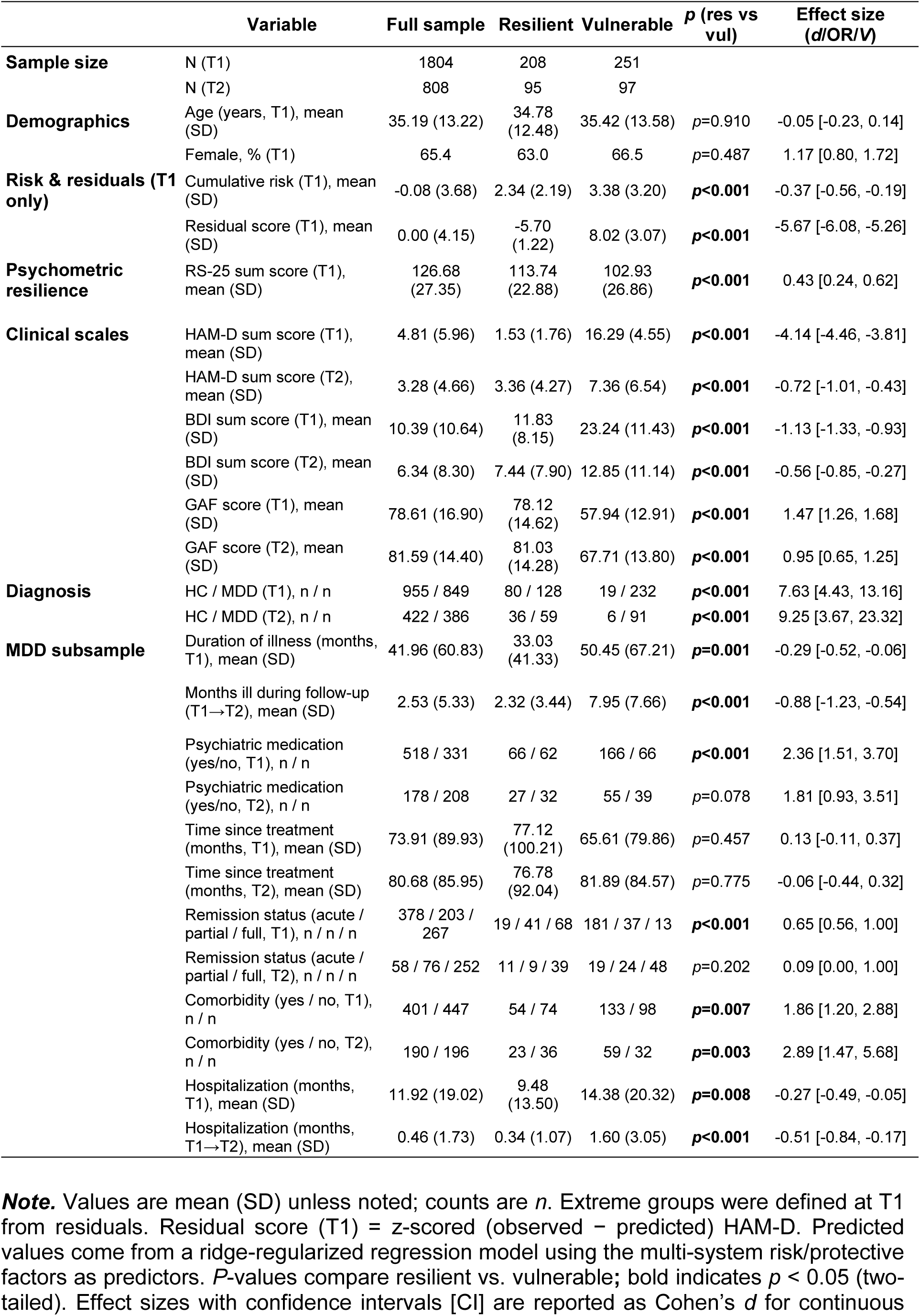
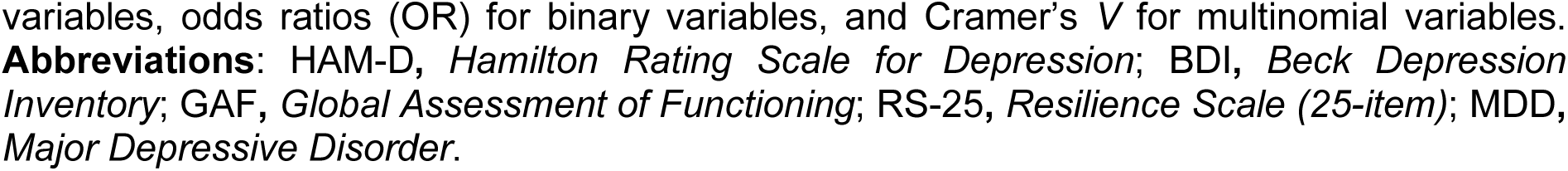
Sample Characteristics.

Sample Characteristics.
|  | Variable | Full sample | Resilient | Vulnerable | *p* (res vs vul) | Effect size (*d*/OR/*V*) |
| --- | --- | --- | --- | --- | --- | --- |
| **Sample size** | N (T1) | 1804 | 208 | 251 |  |  |
|  | N (T2) | 808 | 95 | 97 |  |  |
| **Demographics** | Age (years, T1), mean (SD) | 35.19 (13.22) | 34.78 (12.48) | 35.42 (13.58) | *p*=0.910 | -0.05 [-0.23, 0.14] |
|  | Female, % (T1) | 65.4 | 63.0 | 66.5 | *p*=0.487 | 1.17 [0.80, 1.72] |
| **Risk & residuals (T1 only)** | Cumulative risk (T1), mean (SD) | -0.08 (3.68) | 2.34 (2.19) | 3.38 (3.20) | ***p*<0.001** | -0.37 [-0.56, -0.19] |
|  | Residual score (T1), mean (SD) | 0.00 (4.15) | -5.70 (1.22) | 8.02 (3.07) | ***p*<0.001** | -5.67 [-6.08, -5.26] |
| **Psychometric resilience** | RS-25 sum score (T1), mean (SD) | 126.68 (27.35) | 113.74 (22.88) | 102.93 (26.86) | ***p*<0.001** | 0.43 [0.24, 0.62] |
| **Clinical scales** | HAM-D sum score (T1), mean (SD) | 4.81 (5.96) | 1.53 (1.76) | 16.29 (4.55) | ***p*<0.001** | -4.14 [-4.46, -3.81] |
|  | HAM-D sum score (T2), mean (SD) | 3.28 (4.66) | 3.36 (4.27) | 7.36 (6.54) | ***p*<0.001** | -0.72 [-1.01, -0.43] |
|  | BDI sum score (T1), mean (SD) | 10.39 (10.64) | 11.83 (8.15) | 23.24 (11.43) | ***p*<0.001** | -1.13 [-1.33, -0.93] |
|  | BDI sum score (T2), mean (SD) | 6.34 (8.30) | 7.44 (7.90) | 12.85 (11.14) | ***p*<0.001** | -0.56 [-0.85, -0.27] |
|  | GAF score (T1), mean (SD) | 78.61 (16.90) | 78.12 (14.62) | 57.94 (12.91) | ***p*<0.001** | 1.47 [1.26, 1.68] |
|  | GAF score (T2), mean (SD) | 81.59 (14.40) | 81.03 (14.28) | 67.71 (13.80) | ***p*<0.001** | 0.95 [0.65, 1.25] |
| **Diagnosis** | HC / MDD (T1), *n* / *n* | 955 / 849 | 80 / 128 | 19 / 232 | ***p*<0.001** | 7.63 [4.43, 13.16] |
|  | HC / MDD (T2), *n* / *n* | 422 / 386 | 36 / 59 | 6 / 91 | ***p*<0.001** | 9.25 [3.67, 23.32] |
| **MDD subsample** | Duration of illness (months, T1), mean (SD) | 41.96 (60.83) | 33.03 (41.33) | 50.45 (67.21) | ***p*=0.001** | -0.29 [-0.52, -0.06] |
|  | Months ill during follow-up (T1→T2), mean (SD) | 2.53 (5.33) | 2.32 (3.44) | 7.95 (7.66) | ***p*<0.001** | -0.88 [-1.23, -0.54] |
|  | Psychiatric medication (yes/no, T1), *n* / *n* | 518 / 331 | 66 / 62 | 166 / 66 | ***p*<0.001** | 2.36 [1.51, 3.70] |
|  | Psychiatric medication (yes/no, T2), *n* / *n* | 178 / 208 | 27 / 32 | 55 / 39 | *p*=0.078 | 1.81 [0.93, 3.51] |
|  | Time since treatment (months, T1), mean (SD) | 73.91 (89.93) | 77.12 (100.21) | 65.61 (79.86) | *p*=0.457 | 0.13 [-0.11, 0.37] |
|  | Time since treatment (months, T2), mean (SD) | 80.68 (85.95) | 76.78 (92.04) | 81.89 (84.57) | *p*=0.775 | -0.06 [-0.44, 0.32] |
|  | Remission status (acute / partial / full, T1), *n* / *n* / *n* | 378 / 203 / 267 | 19 / 41 / 68 | 181 / 37 / 13 | ***p*<0.001** | 0.65 [0.56, 1.00] |
|  | Remission status (acute / partial / full, T2), *n* / *n* / *n* | 58 / 76 / 252 | 11 / 9 / 39 | 19 / 24 / 48 | *p*=0.202 | 0.09 [0.00, 1.00] |
|  | Comorbidity (yes / no, T1), *n* / *n* | 401 / 447 | 54 / 74 | 133 / 98 | ***p*=0.007** | 1.86 [1.20, 2.88] |
|  | Comorbidity (yes / no, T2), *n* / *n* | 190 / 196 | 23 / 36 | 59 / 32 | ***p*=0.003** | 2.89 [1.47, 5.68] |
|  | Hospitalization (months, T1), mean (SD) | 11.92 (19.02) | 9.48 (13.50) | 14.38 (20.32) | ***p*=0.008** | -0.27 [-0.49, -0.05] |
|  | Hospitalization (months, T1→T2), mean (SD) | 0.46 (1.73) | 0.34 (1.07) | 1.60 (3.05) | ***p*<0.001** | -0.51 [-0.84, -0.17] |

**Note.** Values are mean (SD) unless noted; counts are *n*. Extreme groups were defined at T1 from residuals. Residual score (T1) = z-scored (observed – predicted) HAM-D. Predicted values come from a ridge-regularized regression model using the multi-system risk/protective factors as predictors. *P*-values compare resilient vs. vulnerable; bold indicates *p* < 0.05 (two-tailed). Effect sizes with confidence intervals [CI] are reported as Cohen's *d* for continuous
variables, odds ratios (OR) for binary variables, and Cramer's $V$ for multinomial variables. **Abbreviations:** HAM-D, *Hamilton Rating Scale for Depression*; BDI, *Beck Depression Inventory*; GAF, *Global Assessment of Functioning*; RS-25, *Resilience Scale (25-item)*; MDD, *Major Depressive Disorder*.

### Structural MRI Acquisition, Preprocessing, and Analyses

MRI acquisition parameter settings and preprocessing steps are detailed in Supplement 2. Analyses followed a hierarchical strategy combining confirmatory and exploratory components to examine associations between residual-based resilience and brain morphometry. Primary (confirmatory) voxel-based morphometry (VBM) region-of-interest (ROI) analyses were conducted based on prior literature implicating specific brain regions in resilience. Statistical significance was assessed using stringent voxel-wise peak-level family-wise error (FWE) correction (α<0.05) within an a priori defined mask (small-volume correction), applying a cluster extent threshold of k≥20 voxels. The combined ROI mask comprised the medial prefrontal cortex (mPFC), orbitofrontal cortex (OFC) and hippocampus. Detailed anatomical definitions and selection rationale are provided in Supplement 3.

Because a priori ROIs were derived from the broader resilience literature rather than from residual-based operationalizations *per se*, complementary whole-brain analyses were conducted to assess the spatial generalizability of observed effects beyond the ROI. These exploratory whole-brain analyses were evaluated using cluster-level FWE correction (α<0.05), with a voxel-wise cluster-defining threshold of p<0.001 (uncorrected) and a minimum cluster extent of k≥20 voxels. For whole-brain analyses, surface-based morphometry (SBM; cortical thickness) was examined alongside GMV.

For both baseline (T1) and two-year follow-up (T2), and across analytic strategies, full-sample regression analyses using T1 residual-based resilience scores as predictors of GMV or cortical thickness were conducted. Analyses examined cross-sectional (T1) and prospective (T1→T2) associations rather than within-subject change over time. These analyses were complemented by extreme-group comparisons based on T1 residual scores (resilient vs. vulnerable) to evaluate robustness of observed associations. Additional sensitivity analyses incorporating cumulative risk, diagnosis or medication as covariates were conducted across strategies and time points to assess stability of results. All statistical contrasts were tested two-sided.

VBM ROI and whole-brain analyses were implemented in SPM using ANCOVA models, specified as multiple regression for full-sample analyses and as full-factorial designs for group comparisons. Covariates included age, sex, and total intracranial volume (TIV) with cumulative risk score, diagnosis, or medication added in additional sensitivity analyses. In accordance with the MRI quality assurance protocol, body- and gradient-coil changes and site were included as additional covariates (77). SBM analyses were conducted using the CAT12 toolbox (CAT12.8.2, r2159), applying the same whole-brain model and covariates as in VBM, omitting TIV. One participant was excluded from SBM analyses at T1 due to corrupted data. For all analyses using T2 data, the interscan interval was included as a covariate. Anatomical labeling for VBM was based on the Neuromorphometrics atlas in Dartel space, and for SBM on the Desikan–Killiany DK40 atlas (78).

## Results

### Sample Characteristics

Table 2 summarizes demographic and clinical characteristics at baseline (T1, N=1,804) and two-year follow-up (T2, N=808) of the full sample and the extreme groups (resilient, vulnerable). Age and sex did not differ between groups. Both extreme groups showed high cumulative risk, with the vulnerable group scoring higher at both time points (*p*<0.001). The resilient group also scored significantly higher on psychometric resilience (RS-25) than the vulnerable group (*p*<0.001). As expected, the vulnerable group showed worse clinical outcomes than the resilient group, including higher HAM-D and Beck’s Depression Inventory (BDI) scores, more comorbidity and lower global assessment of functioning (GAF) scores (all *p*<0.001). From T1 to T2, the vulnerable group spent more months depressed (M=7.95 vs. 2.32; *p*<0.001). Of note, “resilient” does not denote an absence of psychiatric disorder, as 62% of the resilient group met criteria for lifetime MDD at T1.

### Model Performance and Residual-Based Resilience

In the ridge-regularized regression model, cumulative risk explained 51.4% of the variance in depressive symptom severity at T1 (deviance ratio; λ_min_=0.40, MSE=17.01; λ_SE_=4.11, MSE=17.68). Model optimization was based on 10-fold cross-validation, with the final model selected using the 1-SE criterion to enhance generalizability. Cumulative risk strongly predicted HAM-D scores (*r*=0.72, *p*<0.001, see Figure 1). Residual scores, defined as the difference between observed and model-predicted HAM-D scores, index better- or worse-than-predicted outcomes. The relative contribution of predictors is shown in Figure S2. The largest absolute contributions were observed for frequency of social interactions, perceived stress and neuroticism. Correlations among risk/protective factors, residual scores, and HAM-D are shown in Figure S3.

### Temporal Stability of Residual-Based Resilience Over Two Years

To assess stability over the two-year follow-up, we re-estimated the regression model at T2 using the same framework (updating reassessed predictors and carrying forward stable constructs). At T2, the corresponding model yielded a deviance ratio of 44.2% (λ_min_=0.56, MSE=11.84; λ_SE_=5.69, MSE=12.69), and cumulative risk remained a strong predictor for symptom severity (*r*=0.61, *p*<0.001; Figure S1). Residual scores showed moderate temporal stability from T1 to T2 (*r*=0.31, *p*<0.001; Figure S4). The standardized ridge coefficients and correlation structure at T2 are shown in Figures S5 and S6, respectively.

### Cross-Sectional Associations between Residual-Based Resilience and Brain Morphometry

First, we investigated the cross-sectional association between resilience and brain morphometry (GMV and cortical thickness) at T1 in a confirmatory ROI analysis followed by explorative whole-brain analyses. In both the residual-based regression models (GMV: N=1,804; cortical thickness: N=1,803) and the extreme group comparisons (N=459), we found no significant association of resilience with GMV or cortical thickness (see Table S1). This null finding persisted when including cumulative risk or diagnosis or medication as covariate in additional sensitivity analyses (Tables S2-S3). Further, we found no underlying sex by resilience interaction effects (Tables S2-S3).

### Prospective Associations between Residual-Based Resilience and Brain Morphometry

Second, we investigated the prospective (lagged) association between T1 resilience and T2 brain morphometry. Higher residual-based resilience (i.e., larger negative residuals; better-than-expected outcomes) was consistently associated with lower GMV in a cluster comprising the left inferior orbitofrontal gyrus (IOFG) and left temporal pole at two-year follow-up (ROI: *k*=66, *p*_FWE(peak)_=0.002, *r*_partial_=0.18, peak MNI=–48/20/–14; whole-brain: *k*=1572, *p*_FWE(cluster)_=0.008, *r*_partial_=0.18, peak MNI=–50/18/–12; Figure 2, Table 3). This effect emerged in all VBM models (voxel-wise regression, N=808) and in whole-brain extreme group comparisons (N=192, Bonferroni-corrected at *p*<0.025; Figure S7, Table 3). Results remained robust, and in parts slightly strengthened, when adjusting for cumulative risk, diagnosis, or medication (Tables S4-S5). Further, we found no underlying sex by resilience interaction effects (Tables S4-S5). No significant associations were observed for any cortical thickness model of analysis (Table 3; Tables S4-S5).

**Figure 2.**
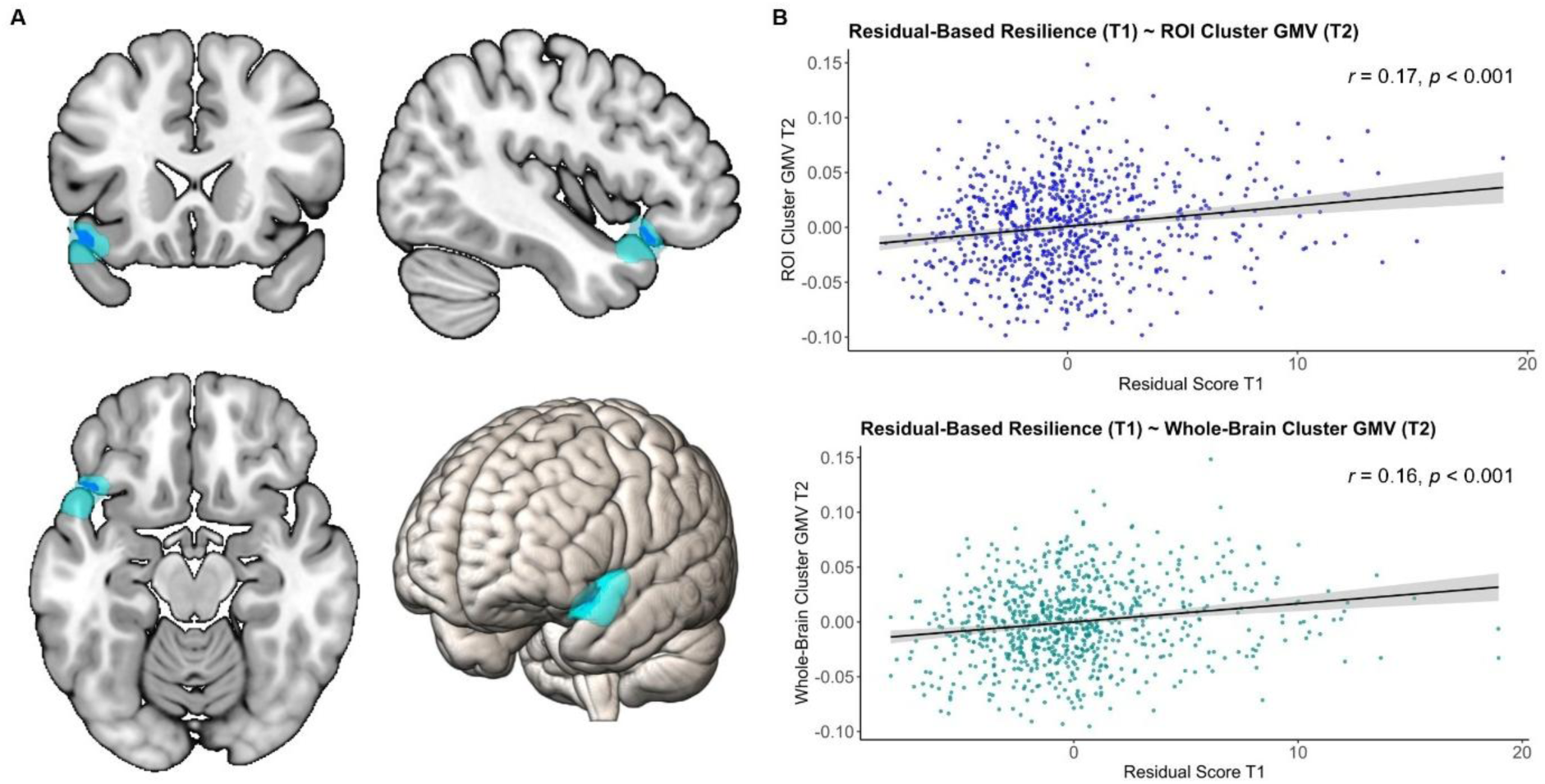
Residual-Based Resilience Predicts Lower GMV at T2 in left IOFG/Temporal Pole. ***Note.*** Regression analyses indicate a robust association between residual-based resilience at T1 (lower residual scores) and lower gray matter volume (GMV) at T2 (first eigenvariate, partial residuals). **A.** Voxels where residual-based resilience at T1 is associated with lower GMV at T2 in regression analyses. ROI-constrained result shown in blue (*k* = 66 voxels, *p*_FWE(peak)_=0.002; *r*_partial_=0.18; MNI=−48/20/-14) and whole-brain result overlayed in cyan (*k*=1572 voxels, *p*_FWE(cluster)_=0.008; *r*_partial_=0.18; MNI=−50/18/−12). **B.** Scatterplots of T1 residual scores versus cluster-mean GMV at T2: ROI cluster *r*=0.17, *p*<0.001; whole-brain cluster *r*=0.16, *p*<0.001. Lower residuals (greater residual-based resilience) correspond to lower GMV.

**Table 3.** Prospective Associations between Residual-Based Resilience (T1) and Brain Morphometry at T2.

| Follow-Up (T2): Residual Score Regression; T1 Resilience → T2 Brain Morphometry (positive association) |  |  |  |  |  |  |  |  |
| --- | --- | --- | --- | --- | --- | --- | --- | --- |
| Modality | Scope | *k* | *p*<sub>FWE(peak)</sub> | *p*<sub>FWE(cluster)</sub> | *T*<sub>800</sub> | Effect Size (partial *r*) | Peak MNI (x/y/z) | Brain Region |
| VBM | ROI | 66 | **0.002** | 0.010 | 5.24 | 0.18 | -48/20/-14 | l. inferior OFG / temporal pole |
| VBM | Whole-Brain | 1572 | 0.002 | **0.008** | 5.28 | 0.18 | -50/18/-12 | l. inferior OFG / temporal pole |
| SBM | Whole-Brain | - | - | - | - | - | - | - |
| Follow-Up (T2): Residual Score Regression; T1 Resilience → T2 Brain Morphometry (negative association) |  |  |  |  |  |  |  |  |
| Modality | Scope | *k* | *p*<sub>FWE(peak)</sub> | *p*<sub>FWE(cluster)</sub> | *T*<sub>800</sub> | Effect Size (partial *r*) | Peak MNI (x/y/z) | Brain Region |
| VBM | ROI | - | - | - | - | - | - | - |
| VBM | Whole-Brain | - | - | - | - | - | - | - |
| SBM | Whole-Brain | 23 | 0.275 | 0.617 | 3.82 | 0.13 | 33/34/28 | r. rostral middle frontal gyrus |
| Follow-Up (T2): Group Comparison; T1 Resilience → T2 Brain Morphometry (vulnerable > resilient) |  |  |  |  |  |  |  |  |
| Modality | Scope | *k* | *p*<sub>FWE(peak)</sub> | *p*<sub>FWE(cluster)</sub> | *T*<sub>184</sub> | Effect Size (Cohen's *d*) | Peak MNI (x/y/z) | Brain Region |
| VBM | ROI | - | - | - | - | - | - | - |
| VBM | Whole-Brain | 1578 | 0.012 | **0.007** | 5.00 | 0.74 | -50/18/-12 | l. inferior OFG / temporal pole |
| SBM | Whole-Brain | - | - | - | - | - | - | - |
| Follow-Up (T2): Group Comparison; T1 Resilience → T2 Brain Morphometry (resilient > vulnerable) |  |  |  |  |  |  |  |  |
| Modality | Scope | *k* | *p*<sub>FWE(peak)</sub> | *p*<sub>FWE(cluster)</sub> | *T*<sub>184</sub> | Effect Size (Cohen's *d*) | Peak MNI (x/y/z) | Brain Region |
| VBM | ROI | - | - | - | - | - | - | - |
| VBM | Whole-Brain | 22 | 0.950 | 0.973 | 3.36 | 0.50 | 60/-33/-24 | r. inferior temporal gyrus |
| SBM | Whole-Brain | - | - | - | - | - | - | - |

**Note.** Summary of voxel-based (VBM) and surface-based (SBM) morphometry results at two-year follow-up (T2). Residual score regression tests the association between T1 residuals and GMV (VBM) or cortical thickness (SBM) at T2. ROI analyses used a priori regions (ACC, subcallosal area, medial frontal gyrus, OFC, hippocampus) with small-volume correction. Group comparisons contrast T1 extreme residual groups (resilient vs. vulnerable); Bonferroni adjustment was applied for the two test directions ( $\alpha=0.025$ ). Effect sizes are partial *r* (regression) and Cohen's *d* (group comparisons). Significant clusters were restricted to the left inferior orbitofrontal gyrus (OFG) / temporal pole. ROI- and whole-brain results refer to the same fronto-temporal effect, identified using different inferential scopes. For analyses without significant effects, the table lists the strongest subthreshold clusters; blank cells indicate that no clusters or peaks emerged even at uncorrected thresholds.

Eigenvariates from the left IOFG/temporal pole cluster at T2 did not correlate with concurrent clinical status or course indicators (HAM-D T2, Social and Occupational Functioning Assessment Scale [SOFAS] T2, months symptom-free, months depressed; all |*r*|<0.05, *p*>0.40; see Figure S8). These null findings suggest that the prospective GMV effect is not simply a proxy for concurrent symptom severity.

## Discussion

The principal finding emerging from this study is that individuals with higher resilience, defined as less depressive symptom severity than expected given their cumulative risk, showed smaller GMV in the left inferior orbitofrontal gyrus and temporal pole at the two-year follow-up, whereas no cross-sectional associations were observed. This pattern suggests that resilience is unlikely to reflect a stable structural feature but rather a delayed neural adaptation within stress-sensitive circuits. Understanding such delayed structural correlates of resilience may help to identify mechanisms that sustain adaptive functioning in the face of adversity.

Applying a dimensional, residual-based approach allowed us to examine resilience beyond diagnostic status and to integrate multiple risk and protective factors across systems. In this resilience framework, the ridge-regularized regression model accounted for more than 50% of the variance in depressive symptom severity at baseline, confirming that cumulative risk robustly explains individual differences in symptom severity. The resulting resilience scores showed moderate stability across the two-year interval, supporting the notion that the residual construct captures both stable, person-specific components and time-varying influences. Conceptually, the moderate stability aligns with a view of resilience as partially enduring yet modifiable over time (1,6,79). Importantly, our residual-based operationalization of resilience does not necessarily equate to resilience as the absence of psychopathology, but rather to a better-than-expected outcome given individual risk exposure. While extremely resilient individuals showed comparatively lower depressive symptom severity, less comorbidity, and better overall functioning than vulnerable individuals, 62% still met criteria for a lifetime MDD diagnosis (47% acute or partially remitted).

At baseline, neither ROI nor whole-brain analyses revealed significant associations between residual-based resilience and brain morphometry. These null results contrast with a number of earlier studies reporting greater GMV in resilient individuals, particularly in prefrontal and limbic regions such as the prefrontal cortex, anterior cingulate cortex, orbitofrontal cortex, and hippocampus (11,13,15), although other studies have reported null or inconsistent findings (16). While differences in sample characteristics likely contribute to heterogeneity across studies (6,10,11,17), methodological variations may likewise account for discrepancies between our findings and prior reports. First, our resilience framework integrates cumulative risk and protective factors across multiple domains, whereas the majority of previous MRI studies examined resilience in response to a single stressor such as childhood trauma. By capturing a broader range of risk factors, this cumulative approach increases ecological validity but may attenuate stressor-specific morphometric effects reported in prior work. Second, most prior studies relied on smaller, diagnostically defined samples, potentially conflating resilience with disorder-related reductions in GMV. Consistent with this notion, several prior reports have described greater or preserved GMV in resilient individuals, notably within regions that are also prone to volume reductions in MDD and PTSD (11,13,16,20–23). Accordingly, a residual-based approach to resilience may help to minimize confounding between resilience-related and disorder-specific sources of variation. Notably, the few studies that have applied such an approach, primarily in adolescent cohorts with conduct disorder, reported only uncorrected or trend-level effects (80,81), suggesting that cross-sectional structural correlates of resilience are subtle and likely context-specific.

In contrast to the absence of baseline associations, higher residual-based resilience at baseline prospectively predicted smaller GMV at T2 in a significant ROI cluster (*k*=66 voxels) located in the left inferior orbitofrontal gyrus and temporal pole, whereas no associations emerged for cortical thickness. This prospective association extended to a larger whole-brain significant cluster (*k*=1,572) with small-to-moderate effect sizes (*r*_partial_=0.18), and was robust across analytic strategies, including whole-brain extreme-group comparisons (medium-to-large effect; *d*=0.74). Importantly, the effect remained significant – and in parts stronger – after adjusting for cumulative risk, clinical diagnosis, and medication, suggesting that it is unlikely to be explained by differences in overall adversity exposure or clinical status. This is particularly relevant as resilience inherently involves exposure to significant adversity, and both adversity (29,33) and psychiatric diagnoses (20,22) have themselves been linked to structural brain alterations. The temporal profile of our findings also accords with evidence that structural adaptations unfold gradually. While baseline contrasts likely capture trait-like predispositions, changes linked to more recent resilience factors may require time to accumulate to a detectable level. Consistent with this view, environmentally induced GMV changes have been demonstrated to emerge over weeks to months in training studies (82,83) and over years under sustained adversity or chronic stress (84–87). No corresponding prospective associations were observed for cortical thickness, suggesting that resilience-related adaptations may be regionally confined to GMV rather than reflecting widespread cortical remodeling.

Both the lateral OFC and the temporal pole are key components of fronto-temporal circuits supporting emotional and social functioning. The left inferior orbitofrontal gyrus, part of the lateral OFC, processes non-reward and punishment stimuli and flexibly updates affective value representations (88–90). The temporal pole, part of the paralimbic system, integrates autobiographical memory, semantic knowledge, and emotional salience to facilitate higher-order social cognition, empathy, and emotion regulation (91–93). Through dense reciprocal connections, the temporal pole provides social and autobiographical context for emotional experiences, while the lateral OFC evaluates their affective value (92,94,95). In depression, this circuit is disrupted, showing weaker connectivity between the medial OFC and temporal pole, which has been linked to reduced retrieval of positive memories, and stronger coupling within the lateral OFC, associated with heightened sensitivity to non-reward and punishment (90,96–98). Within this framework, our finding of smaller GMV in resilient individuals in a cluster encompassing both regions may represent a more selective or efficient configuration of this network, potentially reflecting attenuation of maladaptive over-responsiveness to negative stimuli.

An interesting complementary perspective is offered by the “skin-deep resilience” framework, which suggests that favorable psychological adaptation under adversity may come at biological costs (99–101). Given that resilience presupposes exposure to stress, individuals who adapt well may still experience persistent activation of stress-related biological systems. Under chronic adversity, this sustained activation can lead to allostatic overload, a systemic “wear and tear”, affecting immune, cardiometabolic, and epigenetic processes (85,102–104). In this light, smaller GMV in the IOFG and temporal pole among resilient individuals could also reflect subtle neural costs of exposure to adversity rather than purely adaptive remodeling. The OFC, given its high plasticity and glucocorticoid receptor density, is particularly sensitive to prolonged glucocorticoid signaling and neuroinflammatory processes (105,106). Over time, such mechanisms may induce dendritic remodeling and synaptic loss, contributing to the reduced GMV observed in stress-sensitive cortico-limbic regions (107–110). Taken together, our findings raise the possibility that resilient adaptation in mental health may entail subtle structural costs within fronto-temporal circuits that sustain adaptive functioning under adversity.

Despite the large, well-characterized cohort and comprehensive operationalization of resilience, several limitations of this study should be acknowledged. Although we integrated multiple risk and protective factors across systems, unmeasured influences may still have affected the results. Moreover, current and past depressive episodes can impact brain morphometry and further interact bidirectionally with risk and protective factors (71,111–113). We minimized such confounding by modeling depressive symptom severity as the outcome rather than a predictor and conducting sensitivity analyses controlling for diagnosis, yet some overlap with current symptom status is inherent to residual-based measures. Nevertheless, the absence of typical depression-related morphometric patterns supports the interpretation of resilience-related rather than diagnosis-dependent effects. Our operationalization captures both stable and current risk exposures, implying that long-term effects may have shaped brain structure before baseline, whereas follow-up reflects additional state-dependent influences. Although the prospective design extends beyond prior cross-sectional studies, causal inferences about within-subject structural change remain limited. Moreover, the study lacked biological stress markers (e.g., cortisol), which could have directly tested hypotheses on “wear and tear” or allostatic load. Finally, our analyses were limited to structural MRI. Future work should combine multimodal imaging and physiological markers within longitudinal, residual-based frameworks to clarify whether fronto-temporal alterations reflect adaptive reorganization, biological costs, or both.

Taken together, our findings indicate that resilience to depression is associated with delayed fronto-temporal structural differences that emerge prospectively beyond diagnostic status. These results highlight the importance of considering cumulative risk and dimensional outcomes when investigating neural correlates of resilience.

## Data Availability

All data produced in the present study are available upon reasonable request to the authors / cohort spokepersons

https://zenodo.org/records/17630005

## Acknowledgments and Disclosures

We are deeply indebted to and grateful for all study participants and staff contributing to the MACS cohort. A comprehensive list of acknowledgments can be found at: for2107.de/acknowledgements.

This work is part of the German multicenter consortium “Neurobiology of Affective Disorders. A translational perspective on brain structure and function“, funded by the German Research Foundation (Deutsche Forschungsgemeinschaft DFG; Forschungsgruppe/Research Unit FOR2107). The principal investigators are Tilo Kircher (speaker FOR2107; DFG grant numbers KI 588/14-1, KI 588/14-2, KI 588/15-1, KI 588/17-1), Udo Dannlowski (co-speaker FOR2107; DA 1151/5-1, DA 1151/5-2, DA 1151/6-1, DA1151/9-1, DA1151/10-1, DA1151/11-1), Igor Nenadić (NE 2254/1-2, NE2254/2-1, NE2254/3-1, NE2254/4-1), Markus M. Nöthen (NO 246/10-1, NO 246/10-2), Tim Hahn (HA 7070/2-2), Andreas Jansen (JA 1890/7-1, JA 1890/7-2), Benjamin Straube (STR 1146/18-1). This work was further funded in part by the Germany’s Excellence Strategy (EXC 3066/1 “The Adaptive Mind”, Project No. 533717223). Nina Alexander, Tilo Kircher, Benjamin Straube, and Igor Nenadić were further supported by the Hessian Ministry of Higher Education, Science, Research and Art (LOEWE project “DYNAMIC” Grant Number: LOEWE1/12/519/03/09.001(0009)/98). Udo Dannlowski was additionally funded by the Interdisciplinary Center for Clinical Research (IZKF) of the medical faculty of Münster (grant Dan3/022/22 to UD). This work was supported in part by the SFB/TRR 393 consortium from the German Research Foundation (DFG, project grant no 521379614). The principal investigators are Tilo Kircher (speaker SFB/TRR 393), Nina Alexander, Udo Dannlowski, Tim Hahn, Hamidreza Jamalabadi, Andreas Jansen, Elisabeth J. Leehr, Igor Nenadić, Susanne Meinert, Frederike Stein, and Benjamin Straube.

Tilo Kircher received unrestricted educational grants from Servier, Janssen, Recordati, Aristo, Otsuka, neuraxpharm. The remaining authors have no conflicts of interest.

## CRediT Author Statement

**Conceptualization:** Vincent Hammes, Nina Alexander, Katharina Brosch. **Methodology:** Vincent Hammes, Nina Alexander, Katharina Brosch, Robert Miller, Paula Usemann. **Formal analysis:** Vincent Hammes. **Investigation and Data curation:** Vincent Hammes, Jochen Bauer, Katharina Brosch, Tiana Borgers, Kira Flinkenflügel, Dominik Grotegerd, Tim Hahn, Elisabeth Leehr, Susanne Meinert, Frederike Stein, Albrecht Stroh, Lea Teutenberg, Florian Thomas-Odenthal, Paula Usemann. **Writing – Original Draft:** Vincent Hammes. **Writing – Review & Editing:** Vincent Hammes, Nina Alexander, Jochen Bauer, Katharina Brosch, Tiana Borgers, Udo Dannlowski, Friederike David, Kira Flinkenflügel, Dominik Grotegerd, Tim Hahn, Hamidreza Jamalabadi, Andreas Jansen, Tilo Kircher, Elisabeth Leehr, Susanne Meinert, Robert Miller, Igor Nenadic, Hannah Scherer, Christina Schmitter, Frederike Stein, Benjamin Straube, Albrecht Stroh, Lea Teutenberg, Florian Thomas-Odenthal, Paula Usemann. **Funding acquisition and Resources:** Nina Alexander, Udo Dannlowski, Tim Hahn, Hamidreza Jamalabadi, Andreas Jansen, Tilo Kircher, Frederike Stein, Benjamin Straube. **Supervision:** Nina Alexander.

## Declaration of generative AI and AI-assisted technologies in the writing process

During the preparation of this manuscript, ChatGPT (GPT-5, OpenAI) was used to assist with language editing and refinement. All content was subsequently reviewed and revised by the authors, who take full responsibility for the final version of the manuscript.

