## Supplementary Material for "Resilience Beyond Diagnosis: Prospective Neural Correlates of Better-Than-Expected Outcomes in Depression"

**Hammes *et al.***

**Table of Contents**

**Supplementary Material**

Supplement 1: Risk and Protective Factors in the MACS cohort

Supplement 2: MRI Data Acquisition and Preprocessing

Supplement 3: Region-of Interest Definition and Selection Rationale

**Supplementary Tables**

Supplementary Table 1: Cross-Sectional Associations between Residual-Based Resilience and Brain Morphometry at T1.

Supplementary Table 2: VBM and SBM Brain Morphometry Sensitivity and Interaction Regression Analyses at Baseline (T1).

Supplementary Table 3: VBM and SBM Brain Morphometry Group Comparison Sensitivity and Interaction Analyses at Baseline (T1).

Supplementary Table 4: VBM and SBM Brain Morphometry Sensitivity and Interaction Regression Analyses at Two-Year Follow-Up (T2).

Supplementary Table 5: VBM and SBM Brain Morphometry Group Comparison Sensitivity and Interaction Analyses at Two-Year Follow-Up (T2).

**Supplementary Figures**

Supplementary Figure 1: Ridge-Regularized Regression Model at T2.

Supplementary Figure 2: Impact of Predictor Variables on Outcome at T1.

Supplementary Figure 3: Correlation Matrix of Included Risk and Protective Factors at T1.

Supplementary Figure 4: Stability of Residuals between Baseline (T1) and Two-Year Follow-Up (T2).

Supplementary Figure 5: Impact of Predictor Variables on Outcome at T2.

Supplementary Figure 6: Correlation Matrix of Included Risk and Protective Factors at T2.

Supplementary Figure 7: Resilient Individuals (T1) exhibit Lower GMV at T2 in the left lOFG/Temporal Pole.

Supplementary Figure 8: Associations between Resilience-Associated Cluster Gray Matter Volume and Clinical Course Indicators.

### Supplement 1: Risk and Protective Factors in the MACS cohort

#### Familial Psychiatric Risk

A positive family history of psychiatric disorders is a well-established risk factor for depression. Evidence also suggests transdiagnostic liability across affective and psychotic disorders, indicating overlapping familial risk mechanisms (1,2).

Within the Marburg–Münster Affective Disorders Cohort Study (MACS), familial psychiatric risk was assessed during the socio-demographic interview at baseline (T1) and two-year follow-up (T2). Participants reported whether any first-degree relative (parent, sibling, or child) had a history of Major Depressive Disorder (MDD), Bipolar Disorder (BD), or Schizophrenia (SZ), and whether that relative had received medical or psychological treatment. Two binary indicators were derived: familial risk for affective disorders (AD) and familial risk for psychotic disorders (PD) (coding: 1 = present, 0 = absent). Both variables were entered as dichotomous predictors in the ridge-regularized regression models.

#### Childhood Adversity

Childhood adversity, including childhood maltreatment and adverse family environments, represents a significant and well-established risk factor for MDD and other forms of psychopathology. It has been causally linked to both the development and increased severity of psychiatric disorders, including depression (3–8).

Within the MACS cohort, childhood maltreatment and early adversity were assessed retrospectively using the German versions of the Childhood Trauma Questionnaire (CTQ) (9) and the Adverse Childhood Experiences (ACE) Questionnaire (10). The CTQ was administered at baseline (T1) and at the two-year follow-up (T2) and captures five subtypes of maltreatment (emotional abuse, physical abuse, sexual abuse, emotional neglect, and physical neglect) across 25 items rated on a 5-point Likert scale, with three additional items assessing minimization and denial. Higher scores indicate greater severity or frequency of maltreatment, and the CTQ total sum score was used as a dimensional indicator of overall trauma load. The ACE questionnaire was administered only at baseline (T1) and extends this assessment to ten dichotomous items (yes/no) covering similar domains and additional household dysfunction (i.e., parental separation, domestic violence, substance abuse, mental illness, imprisonment). The ACE sum score reflects cumulative exposure to adverse experiences during childhood and adolescence. Both CTQ and ACE scores were z-standardized within time point and entered the ridge-regression models as continuous predictors.

### Recent Stressful Life Events

Recent stressful life events (SLE) are robustly associated with mental health outcomes and play a crucial role in the onset, course, and recurrence of depression (11–13). However, not all SLE exert detrimental effects as some may also promote adaptive coping or psychological growth through stress inoculation processes (14–17). The subjective evaluation of such events is therefore an important determinant of individual resilience or vulnerability to depression.

Within the MACS cohort, recent SLE experienced within the past six months were assessed at baseline (T1) and SLE experienced since baseline assessment at two-year follow-up (T2) using the German version of the Life Events Questionnaire (LEQ) (18). This self-report instrument captures stressor exposure across major life domains, including health, work, family and friends, finances, and crime. Across 79 items, participants indicated whether specific events occurred and rated (a) their valence (positive/negative) and (b) their subjective impact (0 = no impact to 3 = strong impact). For analysis, negative life event scores were computed by summing the impact ratings of all events that participants had classified as negative. Positive life event scores were calculated analogously for events rated as positive. Higher scores thus represent greater cumulative subjective impact of negative or positive experiences, respectively. Both scores were z-standardized within time point and entered as continuous predictors in the ridge-regression models.

### Perceived Stress

Perceived stress reflects the degree to which situations in one's life are appraised as unpredictable, uncontrollable, or overwhelming. Elevated perceived stress has been consistently associated with increased risk for depression and poorer mental health outcomes (19,20).

Within the MACS cohort, perceived stress during the past month was assessed at both baseline (T1) and two-year follow-up (T2) using the German version of the Perceived Stress Scale (PSS-14) (21). This self-report instrument consists of 14 items evaluating the frequency of stress-related thoughts and feelings on a 5-point Likert scale (0 = never to 4 = very often), with higher scores indicating higher levels of perceived stress. The total PSS sum score was z-standardized within time point and entered as a continuous predictor in the ridge-regression models.

**Social Support**

Social support refers to the perceived availability of social resources and the extent to which individuals are embedded in and connected to a social network. It is a major determinant of mental and physical health, acting as a protective factor against stress and psychopathology (22,23), whereas social isolation and loneliness are established risk factors for adverse health outcomes (24,25).

Within the MACS cohort, perceived social support was assessed at baseline (T1) and two-year follow-up (T2) using the German version of the Perceived Social Support Questionnaire (F-SozU, short form) (26,27). The instrument consists of 22 items rated on a 5-point Likert scale, with higher scores indicating greater perceived social support. The sum score was used as a continuous index of perceived social support. Additionally, a self-constructed item from the socio-demographic questionnaire captured the frequency of current social interactions. Participants indicated how often they had met people outside their family in recent weeks, using a six-point ordinal scale: (1) more than once per week, (2) about once per week, (3) once every two weeks, (4) once per month, (5) none except at work or similar, (6) no interactions under any circumstances. For statistical analyses, the F-SozU sum scores were z-standardized within time point and entered as continuous predictors in the ridge-regression models. The social interaction frequency variable was coded ordinally (1–6, reversed such that higher values indicate less frequent social interaction) and entered as an ordinal predictor.

**Personality**

Personality traits are enduring tendencies to think, feel, and behave in consistent ways across time and situations (28) that are closely associated with mental health outcomes. In particular, high neuroticism has consistently been identified as a major vulnerability factor for depression, whereas extraversion is described as a significant protective factor in mental health (29–32). Conscientiousness and agreeableness have likewise been linked to psychological well-being and positive mental health outcomes (29,33–35). Evidence for openness is more mixed, with some studies reporting associations with cognitive flexibility and well-being (29,36).

Within the MACS cohort, personality traits were assessed using the German version of the Neuroticism–Extraversion–Openness Five-Factor Inventory (NEO-FFI) (37) at baseline (T1). This self-report questionnaire comprises 60 items, with 12 items per trait, rated on a 5-point Likert scale (0 = strongly disagree to 4 = strongly agree). Higher scores indicate stronger expression of the respective trait. Sum scores for each of the five personality dimensions (Neuroticism, Extraversion, Openness, Agreeableness, and Conscientiousness) were z-standardized and entered as continuous predictors in the ridge-regression models.

### Adult Attachment Style

Attachment refers to a relational framework through which individuals form emotional bonds with others, shaping their capacity to engage in close relationships while maintaining autonomy (38). Secure attachment is associated with emotional stability and psychological resilience, whereas insecure attachment constitutes a transdiagnostic risk factor for various mental disorders (39–41).

Within the MACS cohort, adult attachment style was assessed at baseline (T1) using the German version of the Relationship Scales Questionnaire (RSQ) (42,43). The RSQ comprises 30 items rated on a 5-point Likert scale and measures four attachment dimensions: fear of closeness (avoidance), fear of abandonment (anxiety), lack of trust, and independence. Using validated cut-off scores, participants were classified as securely or insecurely attached, with the insecure group further differentiated into fearful, preoccupied, and dismissing attachment styles (42,43). For statistical analyses, attachment style was represented by four binary dummy variables (secure, fearful, preoccupied, and dismissing) in the ridge-regression models (coding: 1 = present, 0 = absent).

### Intelligence

Intelligence reflects general cognitive functioning, encompassing the ability to reason, plan, solve problems, and effectively adapt to the environment (44,45). Cognitive ability has been linked to mental health, where higher intelligence is associated with reduced risk for depression and greater psychological resilience, whereas lower intelligence has been identified as a risk factor for adverse mental health outcomes (32,46–48).

Within the MACS cohort, verbal intelligence was assessed using the Multiple-Choice Vocabulary Test (MWT-B) at both baseline (T1) and two-year follow-up (T2) (49). The MWT-B provides an estimate of crystallized intelligence based on lexical knowledge. Participants are presented with 37 items, each consisting of one real word and four non-words, and asked to identify the correctly spelled word. The total number of correct responses constitutes the MWT-B sum score, which was converted to a verbal intelligence quotient (IQ) estimate using a validated transformation (49). The resulting IQ score was z-standardized within time point, and entered as a continuous predictor in the ridge-regression models.

### Education

Education, as a core indicator of socioeconomic status, is a well-established determinant of mental health. Higher educational attainment is associated with better mental health and well-being, whereas lower education increases the risk for depression and other psychiatric disorders (50–53).

Within the MACS cohort, educational level was assessed at baseline (T1) and two-year follow-up (T2) as part of the socio-demographic questionnaire battery. Participants reported their highest educational qualification, which was converted into total years of education according to the German educational system: compulsory school only (8 years), certificate of secondary education (9 years), general certificate of secondary education (10 years), vocational diploma (12 years), apprenticeship or A levels (13 years), technician (15 years), bachelor's degree (16 years), and master's degree (18 years). Higher academic degrees (e.g., PhD) and interindividual variation in study duration were not considered. For statistical analyses, the years of education were z-standardized within time point and entered as continuous predictors in the ridge-regression models.

**Income**

Socioeconomic conditions such as poverty and wealth are strongly associated with mental health outcomes. Higher income and financial security act as protective factors for psychological well-being, whereas poverty and financial strain increase the risk for both mental and physical health problems (54–57).

Within the MACS cohort, household income was assessed at baseline (T1) and two-year follow-up (T2) as part of the socio-demographic questionnaire battery. Participants provided an estimate of their average monthly net household income, which was capped at 9,999 € to limit outlier influence. Monthly household income was z-standardized and entered as a continuous predictor in the ridge-regression models.

**Immigration**

Immigration constitutes a risk factor for mental illness, a as it often entails exposure to stress, discrimination, social marginalization, and challenges in adapting to a new environment (58,59).

Within the MACS cohort, immigration status was assessed at baseline (T1) as part of the socio-demographic questionnaire battery. Participants were asked whether they or one of their parents had immigrated to Germany from another country. Immigration was recorded as a dichotomous variable (yes/no) and entered as a categorical predictor in the ridge-regression models. It should be noted that all participants are of Central-European ancestry, and thus in this cohort, immigration primarily reflects intra-European migration rather than global migratory diversity.

### Supplement 2: MRI Data Acquisition and Preprocessing

High-resolution T1 images were acquired on 3T MRI scanners (Tim Trio, Siemens, Erlangen, Germany) with 12- and 20- channel head matrix Rx-coils in Marburg and Münster, respectively. A 3D-fast gradient echo sequence (MPRAGE) was used as follows: Marburg: field of view (FOV) = 256 mm, 176 sagittal slices, repetition time (TR) = 1900 ms, echo time (TE) = 2.26 ms, inversion time (TI) = 900 ms, slice thickness = 1 mm, voxel size = 1 × 1 × 1 mm, flip angle = 9°; Münster: FOV = 256 mm, 192 sagittal slices, TR = 2130 ms, TE = 2.28 ms, TI = 900 ms, slice thickness = 1 mm, voxel size = 1 × 1 × 1 mm, flip angle = 8°. Default parameters of the CAT12 toolbox (build 1184, Gaser, Structural Brain Mapping group, Jena University Hospital, Jena, Germany) implemented in SPM12 (v7771, Statistical Parametric Mapping, Institute of Neurology, London, UK) and running under MATLAB (version v2017a, The MathWorks, USA) were used for preprocessing. Images were segmented into gray matter, white matter and cerebrospinal fluid, then normalized using an adapted DARTEL algorithm, and manually quality-checked by a senior clinician. Gaussian kernels with an absolute threshold of 0.1 were used at 8 mm (for GMV) and 20 mm (for cortical thickness) full-width half maximum for data smoothing.

### Supplement 3: Region-of Interest Definition and Selection Rationale

Based on prior work, cortico-limbic regions, particularly the medial prefrontal cortex (mPFC), orbitofrontal cortex (OFC) and hippocampus, are repeatedly implicated in the pathophysiology of and resilience to depression. Functionally, these regions support adaptive emotion regulation and stress recovery, providing the mechanistic rationale for their a priori selection (60–66). Using the Neuromorphometrics atlas in DARTEL-space (SPM12), we defined a bilateral region-of-interest (ROI) mask comprising the mPFC (superior medial frontal gyri, anterior cingulate gyri, and subcallosal area) and the OFC (anterior, medial, lateral, and posterior orbital gyri and the inferior frontal orbital gyri), together with the bilateral hippocampi. This bilateral mask was used for ROI analyses, whereas whole-brain analyses imposed no regional restrictions. Most structural resilience studies have used VBM. By contrast, surface-based measures such as cortical thickness, with partly distinct genetic determinants, complement GMV and add nonredundant information (67,68). Given limited prior cortical-thickness findings in resilience, SBM analyses were treated as exploratory and conducted at the whole-brain level.

### Supplementary Material – Tables

#### Supplementary Table 1.

Cross-Sectional Associations between Residual-Based Resilience and Brain Morphometry at T1.

**Baseline (T1): Residual Score Regression (positive association)**

| Modality | Scope | *k* | $p_{FWE(peak)}$ | $p_{FWE(cluster)}$ | $T_{1796}$ | Effect Size (partial *r*) | Peak MNI (*x/y/z*) | Brain Region |
|---|---|---|---|---|---|---|---|---|
| VBM | ROI | - | - | - | - | - | - | - |
| VBM | Whole-Brain | - | - | - | - | - | - | - |
| SBM | Whole-Brain | - | - | - | - | - | - | - |

**Baseline (T1): Residual Score Regression (negative association)**

| Modality | Scope | *k* | $p_{FWE(peak)}$ | $p_{FWE(cluster)}$ | $T_{1796}$ | Effect Size (partial *r*) | Peak MNI (*x/y/z*) | Brain Region |
|---|---|---|---|---|---|---|---|---|
| VBM | ROI | - | - | - | - | - | - | - |
| VBM | Whole-Brain | 140 | 0.518 | 0.748 | 3.76 | 0.09 | 33/-18/-20 | r. hippocampus |
| SBM | Whole-Brain | 39 | 0.132 | 0.421 | 3.75 | 0.09 | 54/-34/-20 | r. inferior temporal gyrus |

**Baseline (T1): Group Comparison (resilient > vulnerable)**

| Modality | Scope | *k* | $p_{FWE(peak)}$ | $p_{FWE(cluster)}$ | $T_{449}$ | Effect Size (Cohen's *d*) | Peak MNI (*x/y/z*) | Brain Region |
|---|---|---|---|---|---|---|---|---|
| VBM | ROI | - | - | - | - | - | - | - |
| VBM | Whole-Brain | 39 | 0.541 | 0.950 | 3.77 | 0.36 | -15/40/-12 | l. medial orbital gyrus |
| SBM | Whole-Brain | - | - | - | - | - | - | - |

**Baseline (T1): Group Comparison (vulnerable > resilient)**

| Modality | Scope | *k* | $p_{FWE(peak)}$ | $p_{FWE(cluster)}$ | $T_{449}$ | Effect Size (Cohen's *d*) | Peak MNI (*x/y/z*) | Brain Region |
|---|---|---|---|---|---|---|---|---|
| VBM | ROI | - | - | - | - | - | - | - |
| VBM | Whole-Brain | 46 | 0.760 | 0.940 | 3.58 | 0.34 | 10/-30/16 | r. thalamus proper |
| SBM | Whole-Brain | - | - | - | - | - | - | - |

***Note.*** Summary of voxel-based (VBM) and surface-based (SBM) morphometry results at baseline (T1). Residual score regression tests the association between T1 residuals and GMV (VBM) or cortical thickness (SBM) at T1. ROI analyses used a priori regions (ACC, subcallosal area, medial frontal gyrus, OFC, hippocampus) with small-volume correction. Group comparisons contrast T1 extreme residual groups (resilient vs. vulnerable); Bonferroni adjustment was applied for the two test directions ($\alpha$ = 0.025). Effect sizes are partial *r* (regression) and Cohen's *d* (group comparisons). As no significant associations emerged, the table lists the strongest subthreshold clusters; blank cells indicate that no clusters or peaks were detected even at uncorrected thresholds. Importantly, none of the subthreshold clusters were consistent across analytic approaches (ROI vs. whole-brain), modalities (VBM vs. SBM), or sensitivity models, and therefore were not interpreted further.

**Supplementary Table 2.**

VBM and SBM Brain Morphometry Sensitivity and Interaction Regression Analyses at Baseline (T1).

**Baseline (T1): Residual Score Regression (positive association)**

| **VBM Model** | ***k*** | $p_{FWE(peak)}$ | $p_{FWE(cluster)}$ | $T_{1795}$ | **Effect Size (partial *r*)** | **Peak MNI (*x/y/z*)** | **Brain Region** |
|---|---|---|---|---|---|---|---|
| ROI: Sensitivity Model Cumulative Risk | - | - | - | - | - | - | - |
| ROI: Sensitivity Model Diagnosis | - | - | - | - | - | - | - |
| ROI: Sensitivity Model Medication | - | - | - | - | - | - | - |
| ROI: Interaction (Sex by Resilience) | - | - | - | - | - | - | - |
| Whole-Brain: Sensitivity Model Cumulative Risk | - | - | - | - | - | - | - |
| Whole-Brain: Sensitivity Model Diagnosis | - | - | - | - | - | - | - |
| Whole-Brain: Sensitivity Model Medication | - | - | - | - | - | - | - |
| Whole-Brain: Interaction (Sex by Resilience) | 57 | 0.934 | 0.914 | 3.32 | 0.08 | -60/-16/33 | l. postcentral gyrus |
| **SBM Model** | ***k*** | $p_{FWE(peak)}$ | $p_{FWE(cluster)}$ | $T_{1795}$ | **Effect Size (partial *r*)** | **Peak MNI (*x/y/z*)** | **Brain Region** |
| Whole-Brain: Sensitivity Model Cumulative Risk | - | - | - | - | - | - | - |
| Whole-Brain: Sensitivity Model Diagnosis | - | - | - | - | - | - | - |
| Whole-Brain: Sensitivity Model Medication | - | - | - | - | - | - | - |
| Whole-Brain: Interaction (Sex by Resilience) | - | - | - | - | - | - | - |

**Baseline (T1): Residual Score Regression (negative association)**

| **VBM Model** | ***k*** | $p_{FWE(peak)}$ | $p_{FWE(cluster)}$ | $T_{1795}$ | **Effect Size (partial *r*)** | **Peak MNI (*x/y/z*)** | **Brain Region** |
|---|---|---|---|---|---|---|---|
| ROI: Sensitivity Model Cumulative Risk | - | - | - | - | - | - | - |
| ROI: Sensitivity Model Diagnosis | - | - | - | - | - | - | - |
| ROI: Sensitivity Model Medication | - | - | - | - | - | - | - |
| ROI: Interaction (Sex by Resilience) | - | - | - | - | - | - | - |
| Whole-Brain: Sensitivity Model Cumulative Risk | 63 | 0.795 | 0.904 | 3.51 | 0.08 | 33/-16/-21 | r. hippocampus |
| Whole-Brain: Sensitivity Model Diagnosis | 70 | 0.778 | 0.891 | 3.53 | 0.08 | -40/26/14 | l. inferior frontal angular gyrus |
| Whole-Brain: Sensitivity Model Medication | 56 | 0.824 | 0.916 | 3.48 | 0.08 | -40/26/14 | l. inferior frontal angular gyrus |
| Whole-Brain: Interaction (Sex by Resilience) | 188 | 0.021 | 0.648 | 4.70 | 0.11 | -04/-39/08 | l. posterior cingulate gyrus |
| **SBM Model** | ***k*** | $p_{FWE(peak)}$ | $p_{FWE(cluster)}$ | $T_{1795}$ | **Effect Size (partial *r*)** | **Peak MNI (*x/y/z*)** | **Brain Region** |
| Whole-Brain: Sensitivity Model Cumulative Risk | - | - | - | - | - | - | - |
| Whole-Brain: Sensitivity Model Diagnosis | - | - | - | - | - | - | - |
| Whole-Brain: Sensitivity Model Medication | - | - | - | - | - | - | - |
| Whole-Brain: Interaction (Sex by Resilience) | - | - | - | - | - | - | - |

***Note.*** Summary of voxel-based (VBM) and surface-based (SBM) sensitivity and interaction regression analyses at baseline (T1). Residual score regression tests the association between T1 residuals and GMV (VBM) or cortical thickness (SBM) at T1. ROI analyses used a priori regions (ACC, subcallosal area, medial frontal gyrus, OFC, hippocampus) with small-volume correction. Sensitivity models included cumulative risk, clinical diagnosis, or medication as additional covariates; interaction models tested the sex-by-residual interaction. Effect sizes are partial *r*. As no significant associations emerged, the table lists the strongest subthreshold clusters; blank cells indicate that no clusters or peaks were detected even at uncorrected thresholds. Importantly, none of the subthreshold clusters were consistent across analytic approaches (ROI vs. whole-brain), modalities (VBM vs. SBM), or sensitivity models, and therefore were not interpreted further.

**Supplementary Table 3.**

VBM and SBM Brain Morphometry Group Comparison Sensitivity and Interaction Analyses at Baseline (T1).

**Baseline (T1): Group Comparison**

| **VBM Model** | ***k*** | $p_{FWE(peak)}$ | $p_{FWE(cluster)}$ | $T_{448}$ | **Effect Size (Cohen's *d*)** | **Peak MNI (*x/y/z*)** | **Brain Region** |
|---|---|---|---|---|---|---|---|
| ROI: Sensitivity Model Cumulative Risk | | | | | | | |
| *resilient* > *vulnerable* | - | - | - | - | - | - | - |
| *vulnerable*> *resilient* | - | - | - | - | - | - | - |
| ROI: Sensitivity Model Diagnosis | | | | | | | |
| *resilient* > *vulnerable* | - | - | - | - | - | - | - |
| *vulnerable*> *resilient* | - | - | - | - | - | - | - |
| ROI: Sensitivity Model Medication | | | | | | | |
| *resilient* > *vulnerable* | - | - | - | - | - | - | - |
| *vulnerable*> *resilient* | - | - | - | - | - | - | - |
| Whole-Brain: Sensitivity Model Cumulative Risk | | | | | | | |
| *resilient* > *vulnerable* | 31 | 0.696 | 0.962 | 3.64 | 0.34 | -15/40/-12 | l. medial orbital gyrus |
| *vulnerable*> *resilient* | 41 | 0.786 | 0.948 | 3.55 | 0.34 | 10/-30/16 | r. thalamus proper |
| Whole-Brain: Sensitivity Model Diagnosis | | | | | | | |
| *resilient* > *vulnerable* | 163 | 0.244 | 0.704 | 4.08 | 0.39 | -15/40/-14 | l. medial orbital gyrus |
| *vulnerable*> *resilient* | 50 | 0.715 | 0.934 | 3.62 | 0.34 | 09/-28/16 | r. thalamus proper |
| Whole-Brain: Sensitivity Model Medication | | | | | | | |
| *resilient* > *vulnerable* | 34 | 0.663 | 0.957 | 3.67 | 0.35 | -15/40/-12 | l. medial orbital gyrus |
| *vulnerable*> *resilient* | 85 | 0.345 | 0.870 | 3.96 | 0.37 | 10/-30/16 | r. thalamus proper |
| Whole-Brain: Interaction (Group by Sex) | | | | | | | |
| *positive interaction* | - | - | - | - | - | - | - |
| *negative interaction* | 162 | 0.020 | 0.708 | 4.77 | 0.45 | -04/-39/06 | l. posterior cingulate gyrus |
| **SBM Model** | ***k*** | $p_{FWE(peak)}$ | $p_{FWE(cluster)}$ | $T_{448}$ | **Effect Size (Cohen's *d*)** | **Peak MNI (*x/y/z*)** | **Brain Region** |
| Whole-Brain: Sensitivity Model Cumulative Risk | | | | | | | |
| *resilient* > *vulnerable* | - | - | - | - | - | - | - |
| *vulnerable*> *resilient* | - | - | - | - | - | - | - |
| Whole-Brain: Sensitivity Model Diagnosis | | | | | | | |
| *resilient* > *vulnerable* | - | - | - | - | - | - | - |
| *vulnerable*> *resilient* | - | - | - | - | - | - | - |
| Whole-Brain: Sensitivity Model Medication | | | | | | | |
| *resilient* > *vulnerable* | - | - | - | - | - | - | - |
| *vulnerable*> *resilient* | - | - | - | - | - | - | - |
| Whole-Brain: Interaction (Group by Sex) | | | | | | | |
| *positive interaction* | - | - | - | - | - | - | - |
| *negative interaction* | - | - | - | - | - | - | - |

***Note.*** Summary of voxel-based (VBM) and surface-based (SBM) group comparison sensitivity and interaction analyses at baseline (T1). Group comparisons contrast T1 extreme residual groups (resilient vs. vulnerable) for GMV (VBM) or cortical thickness (SBM). Bonferroni adjustment was applied for the two test directions ($\alpha$ = 0.025). ROI analyses used a priori regions (ACC, subcallosal area, medial frontal gyrus, OFC, hippocampus) with small-volume correction. Sensitivity models included cumulative risk, clinical diagnosis, or medication as additional covariates; interaction models tested the group-by-sex interaction. Effect sizes are Cohen's *d*. As no significant associations emerged, the table lists the strongest subthreshold clusters; blank cells indicate that no clusters or peaks were detected even at uncorrected thresholds. Importantly, none of the subthreshold clusters were consistent across analytic approaches (ROI vs. whole-brain), modalities (VBM vs. SBM), or sensitivity models, and therefore were not interpreted further.

**Supplementary Table 4.**

VBM and SBM Brain Morphometry Sensitivity and Interaction Regression Analyses at Two-Year Follow-Up (T2).

**Follow-Up (T2): Residual Score Regression; T1 Resilience → T2 Brain Morphometry (positive association)**

| **VBM Model** | ***k*** | ***p*$_{FWE(peak)}$** | ***p*$_{FWE(cluster)}$** | ***T*$_{799}$** | **Effect Size (partial *r*)** | **Peak MNI (*x/y/z*)** | **Brain Region** |
|---|---|---|---|---|---|---|---|
| ROI: Sensitivity Model Cumulative Risk | 100 | **<0.001** | 0.006 | 5.60 | 0.19 | -48/20/-14 | l. inferior OFG / temporal pole |
| ROI: Sensitivity Model Diagnosis | 79 | **0.001** | 0.008 | 5.40 | 0.19 | -48/20/-14 | l. inferior OFG / temporal pole |
| ROI: Sensitivity Model Medication | 137 | **<0.001** | 0.003 | 5.85 | 0.20 | -48/20/-14 | l. inferior OFG / temporal pole |
| ROI: Interaction (Sex by Resilience) | - | - | - | - | - | - | |
| Whole-Brain: Sensitivity Model Cumulative Risk | 1652 | 0.001 | **0.007** | 5.40 | 0.19 | -48/18/-12 | l. inferior OFG / temporal pole |
| Whole-Brain: Sensitivity Model Diagnosis | 1844 | <0.001 | **0.004** | 5.62 | 0.20 | -48/18/-12 | l. inferior OFG / temporal pole |
| Whole-Brain: Sensitivity Model Medication | 2059 | <0.001 | **0.002** | 5.85 | 0.20 | -48/20/-14 | l. inferior OFG / temporal pole |
| Whole-Brain: Interaction (Sex by Resilience) | - | - | - | - | - | - | - |

| **SBM Model** | ***k*** | ***p*$_{FWE(peak)}$** | ***p*$_{FWE(cluster)}$** | ***T*$_{799}$** | **Effect Size (partial *r*)** | **Peak MNI (*x/y/z*)** | **Brain Region** |
|---|---|---|---|---|---|---|---|
| Whole-Brain: Sensitivity Model Cumulative Risk | - | - | - | - | - | - | - |
| Whole-Brain: Sensitivity Model Diagnosis | - | - | - | - | - | - | - |
| Whole-Brain: Sensitivity Model Medication | - | - | - | - | - | - | - |
| Whole-Brain: Interaction (Sex by Resilience) | - | - | - | - | - | - | - |

**Follow-Up (T2): Residual Score Regression; T1 Resilience → T2 Brain Morphometry (negative association)**

| **VBM Model** | ***k*** | ***p*$_{FWE(peak)}$** | ***p*$_{FWE(cluster)}$** | ***T*$_{799}$** | **Effect Size (partial *r*)** | **Peak MNI (*x/y/z*)** | **Brain Region** |
|---|---|---|---|---|---|---|---|
| ROI: Sensitivity Model Cumulative Risk | - | - | - | - | - | - | - |
| ROI: Sensitivity Model Diagnosis | - | - | - | - | - | - | - |
| ROI: Sensitivity Model Medication | - | - | - | - | - | - | - |
| ROI: Interaction (Sex by Resilience) | - | - | - | - | - | - | - |
| Whole-Brain: Sensitivity Model Cumulative Risk | - | - | - | - | - | - | - |
| Whole-Brain: Sensitivity Model Diagnosis | 21 | 0.904 | 0.969 | 3.38 | 0.12 | -02/08/04 | l. caudate |
| Whole-Brain: Sensitivity Model Medication | - | - | - | - | - | - | - |
| Whole-Brain: Interaction (Sex by Resilience) | 108 | 0.516 | 0.816 | 3.77 | 0.13 | 26/27/36 | r. middle frontal gyrus |

| **SBM Model** | ***k*** | ***p*$_{FWE(peak)}$** | ***p*$_{FWE(cluster)}$** | ***T*$_{799}$** | **Effect Size (partial *r*)** | **Peak MNI (*x/y/z*)** | **Brain Region** |
|---|---|---|---|---|---|---|---|
| Whole-Brain: Sensitivity Model Cumulative Risk | 30 | 0.165 | 0.492 | 3.98 | 0.14 | 33/34/28 | r. rostral middle frontal gyrus |
| Whole-Brain: Sensitivity Model Diagnosis | 45 | 0.033 | 0.276 | 4.41 | 0.15 | 33/34/28 | r. rostral middle frontal gyrus |
| Whole-Brain: Sensitivity Model Medication | 23 | 0.161 | 0.617 | 3.99 | 0.14 | 54/-34/-21 | r. inferior temporal gyrus |
| Whole-Brain: Interaction (Sex by Resilience) | - | - | - | - | - | - | - |

***Note.*** Summary of voxel-based (VBM) and surface-based (SBM) sensitivity and interaction regression analyses at two-year follow-up (T2). Residual score regression tests the association between T1 residuals and GMV (VBM) or cortical thickness (SBM) at T2. ROI analyses used a priori regions (ACC, subcallosal area, medial frontal gyrus, OFC, hippocampus) with small-volume correction. Sensitivity models included cumulative risk, clinical diagnosis, or medication as additional covariates; interaction models tested the residual-by-sex interaction. Effect sizes are partial *r*. Significant clusters were confined to the left inferior orbitofrontal gyrus (OFG) / temporal pole region. ROI- and whole-brain significant results refer to the same fronto-temporal effect, identified using different inferential scopes. For models without significant associations, the table lists the strongest subthreshold clusters; blank cells indicate that no clusters or peaks were detected even at uncorrected thresholds.

**Supplementary Table 5.**

VBM and SBM Brain Morphometry Group Comparison Sensitivity and Interaction Analyses at Two-Year Follow-Up (T2).

**Follow-Up (T2): Group Comparison; T1 Resilience → T2 Brain Morphometry**

| **VBM Model** | ***k*** | $p_{FWE(peak)}$ | $p_{FWE(cluster)}$ | $T_{183}$ | **Effect Size (Cohen's *d*)** | **Peak MNI (*x/y/z*)** | **Brain Region** |
|---|---|---|---|---|---|---|---|
| ROI: Sensitivity Model Cumulative Risk | | | | | | | |
| *resilient > vulnerable* | - | - | - | - | - | - | - |
| *vulnerable > resilient* | - | - | - | - | - | - | - |
| ROI: Sensitivity Model Diagnosis | | | | | | | |
| *resilient > vulnerable* | - | - | - | - | - | - | - |
| *vulnerable > resilient* | - | - | - | - | - | - | - |
| ROI: Sensitivity Model Medication | | | | | | | |
| *resilient > vulnerable* | - | - | - | - | - | - | - |
| *vulnerable > resilient* | - | - | - | - | - | - | - |
| ROI: Interaction (Group by Sex) | | | | | | | |
| *Positive Interaction* | - | - | - | - | - | - | - |
| *Negative Interaction* | - | - | - | - | - | - | - |
| Whole-Brain: Sensitivity Model Cumulative Risk | | | | | | | |
| *resilient > vulnerable* | 29 | 0.924 | 0.965 | 3.36 | 0.50 | -10/44/24 | l. superior medial frontal gyrus |
| *vulnerable > resilient* | 1555 | 0.015 | **0.008** | 4.95 | 0.73 | -51/18/-12 | l. inferior OFG / temporal pole |
| Whole-Brain: Sensitivity Model Diagnosis | | | | | | | |
| *resilient > vulnerable* | 65 | 0.808 | 0.910 | 3.58 | 0.53 | -10/42/24 | l. superior medial frontal gyrus |
| *vulnerable > resilient* | 1405 | 0.034 | **0.012** | 4.73 | 0.70 | -50/18/-12 | l. inferior OFG / temporal pole |
| Whole-Brain: Sensitivity Model Medication | | | | | | | |
| *resilient > vulnerable* | - | - | - | - | - | - | |
| *vulnerable > resilient* | 1877 | 0.002 | **0.003** | 5.48 | 0.81 | -50/18/-12 | l. inferior OFG / temporal pole |
| Whole-Brain: Interaction (Group by Sex) | | | | | | | |
| *Positive Interaction* | - | - | - | - | - | - | |
| *Negative Interaction* | 52 | 0.494 | 0.938 | 3.90 | 0.58 | -04/-33/04 | l. thalamus proper |

| **SBM Model** | ***k*** | $p_{FWE(peak)}$ | $p_{FWE(cluster)}$ | $T_{183}$ | **Effect Size (Cohen's *d*)** | **Peak MNI (*x/y/z*)** | **Brain Region** |
|---|---|---|---|---|---|---|---|
| Whole-Brain: Sensitivity Model Cumulative Risk | | | | | | | |
| *resilient > vulnerable* | - | - | - | - | - | - | - |
| *vulnerable > resilient* | - | - | - | - | - | - | - |
| Whole-Brain: Sensitivity Model Diagnosis | | | | | | | |
| *resilient > vulnerable* | - | - | - | - | - | - | - |
| *vulnerable > resilient* | - | - | - | - | - | - | - |
| Whole-Brain: Sensitivity Model Medication | | | | | | | |
| *resilient > vulnerable* | - | - | - | - | - | - | - |
| *vulnerable > resilient* | - | - | - | - | - | - | - |
| Whole-Brain: Interaction (Group by Sex) | | | | | | | |
| *Positive Interaction* | - | - | - | - | - | - | - |
| *Negative Interaction* | - | - | - | - | - | - | - |

***Note.*** Summary of voxel-based (VBM) and surface-based (SBM) group comparison sensitivity and interaction analyses at two-year follow-up (T2). Group comparisons contrast T1 extreme residual groups (resilient vs. vulnerable) for GMV (VBM) or cortical thickness (SBM) at T2. Bonferroni adjustment was applied for the two test directions ($\alpha$ = 0.025). ROI analyses used a priori regions (ACC, subcallosal area, medial frontal gyrus, OFC, hippocampus) with small-volume correction. Sensitivity models included cumulative risk, clinical diagnosis, or medication as additional covariates; interaction models tested the group-by-sex interaction. Effect sizes are Cohen's *d*. Significant clusters were confined to the left inferior orbitofrontal gyrus (OFG) / temporal pole region. ROI- and whole-brain results refer to the same fronto-temporal effect, identified using different inferential scopes. For analyses without significant associations, the table lists the strongest subthreshold clusters; blank cells indicate that no clusters or peaks were detected even at uncorrected thresholds.

**Supplementary Material – Figures**

**Supplementary Figure 1.** Ridge-Regularized Regression Model at T2.

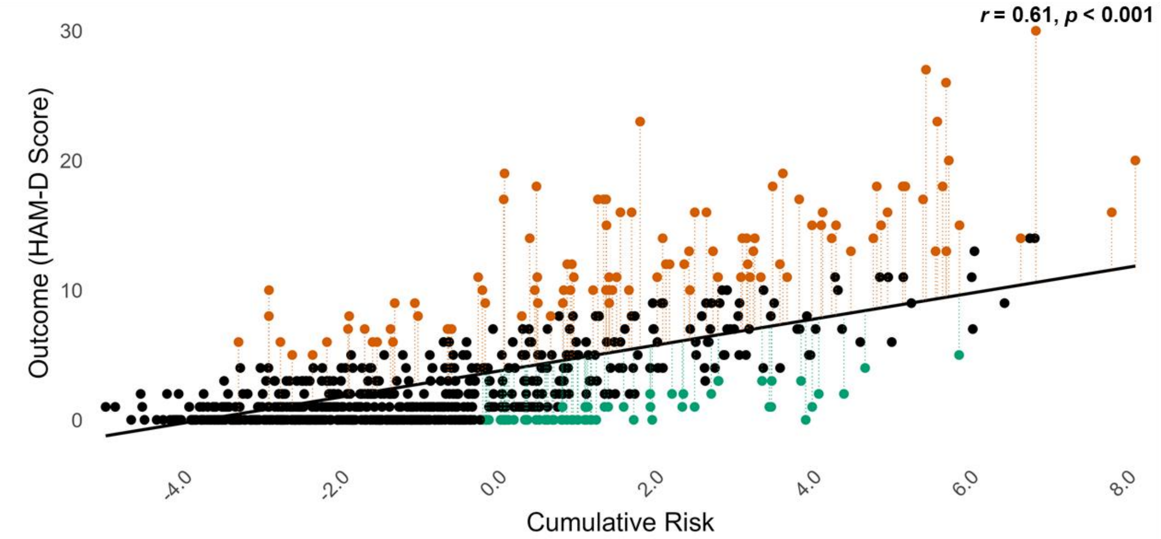


***Note.*** Ridge regression at T2 showing the significant positive association between cumulative risk and depressive symptom severity (HAM-D, $r = 0.61$, $p < 0.001$). Residuals reflect individual deviations from predicted symptom scores and index resilience (green; better-than-expected) and vulnerability (red; worse-than-expected), defined as values exceeding ±1 residual standard error (RSE = 3.58).

**Supplementary Figure 2.** Impact of Predictor Variables on Outcome at T1.

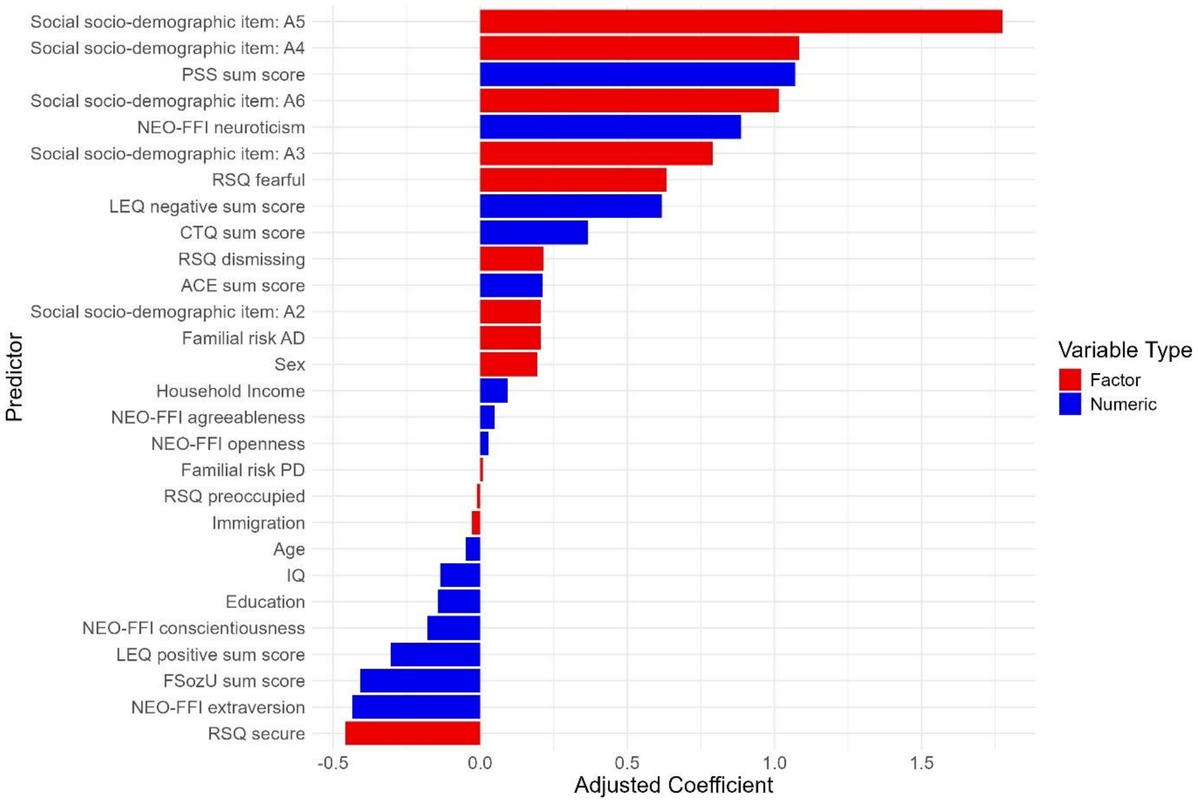


***Note***. Standardized (blue) and unstandardized (red) regression coefficients of all predictors at T1. For continuous predictors, coefficients were scaled by the predictor's standard deviation (SD) and represent the expected change in HAM-D sum scores per 1-SD increase. Positive coefficients indicate higher predicted depressive symptom severity. Categorical predictors are shown as unstandardized coefficients and reflect differences relative to the reference category (e.g., attachment style present vs. absent; familial risk vs. none; female vs. male sex). The ordinal scaled social interaction answer items A2-A6 (increasingly fewer social interactions) reflect differences relative to A1 ("more than one social interaction per week").

**Supplementary Figure 3.** Correlation Matrix of Included Risk and Protective Factors at T1.

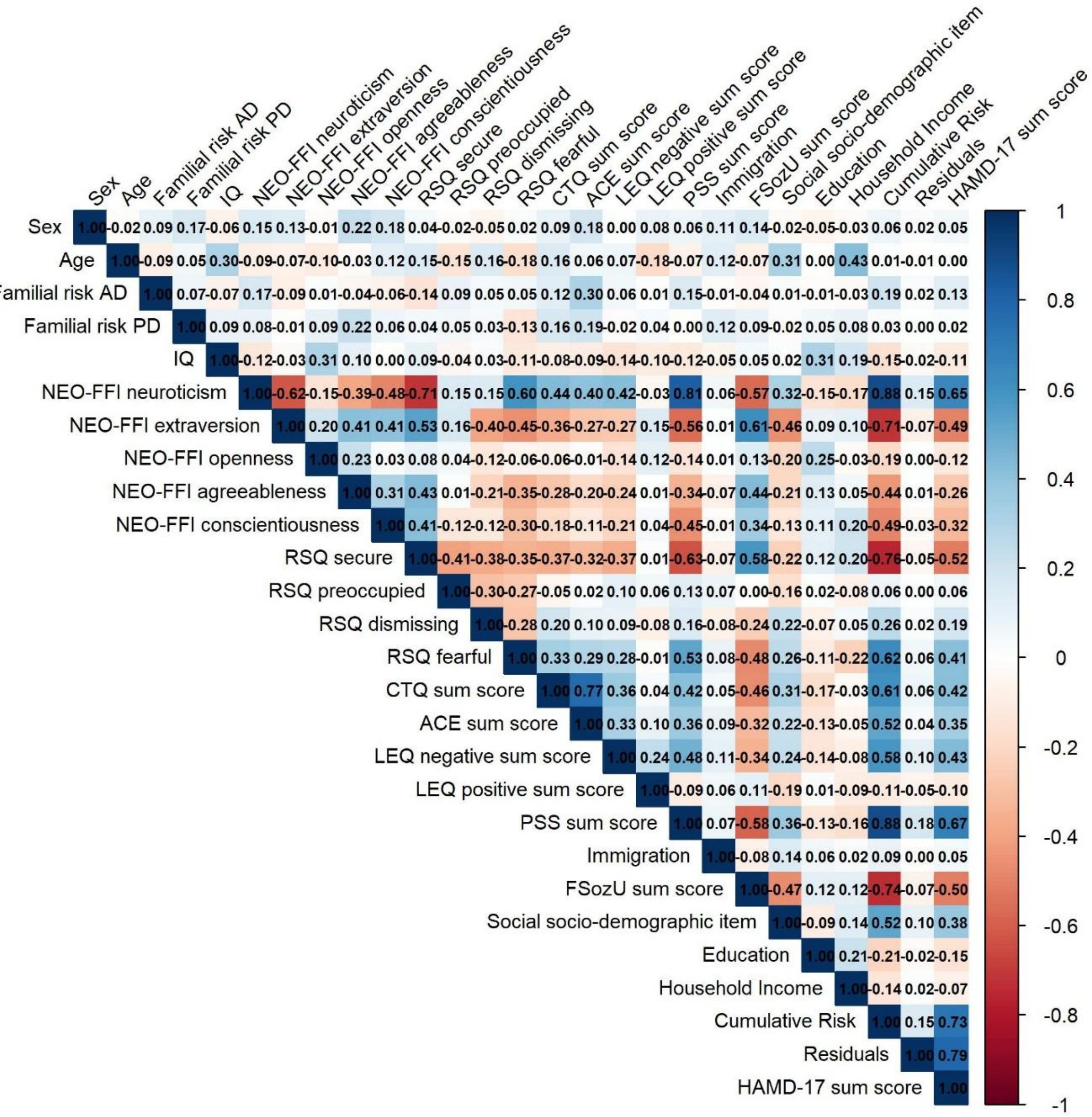


***Note***. Correlation matrix of all predictors at T1, including the 22 risk and protective factors, age, sex, cumulative risk, residual scores, and HAM-D. Pearson correlations were computed for continuous variable pairs, polychoric correlations for categorical pairs, and polyserial correlations for mixed continuous–categorical pairs. Values represent correlation coefficients (*r*).

**Supplementary Figure 4.** Stability of Residuals between Baseline (T1) and Two-Year Follow-Up (T2).

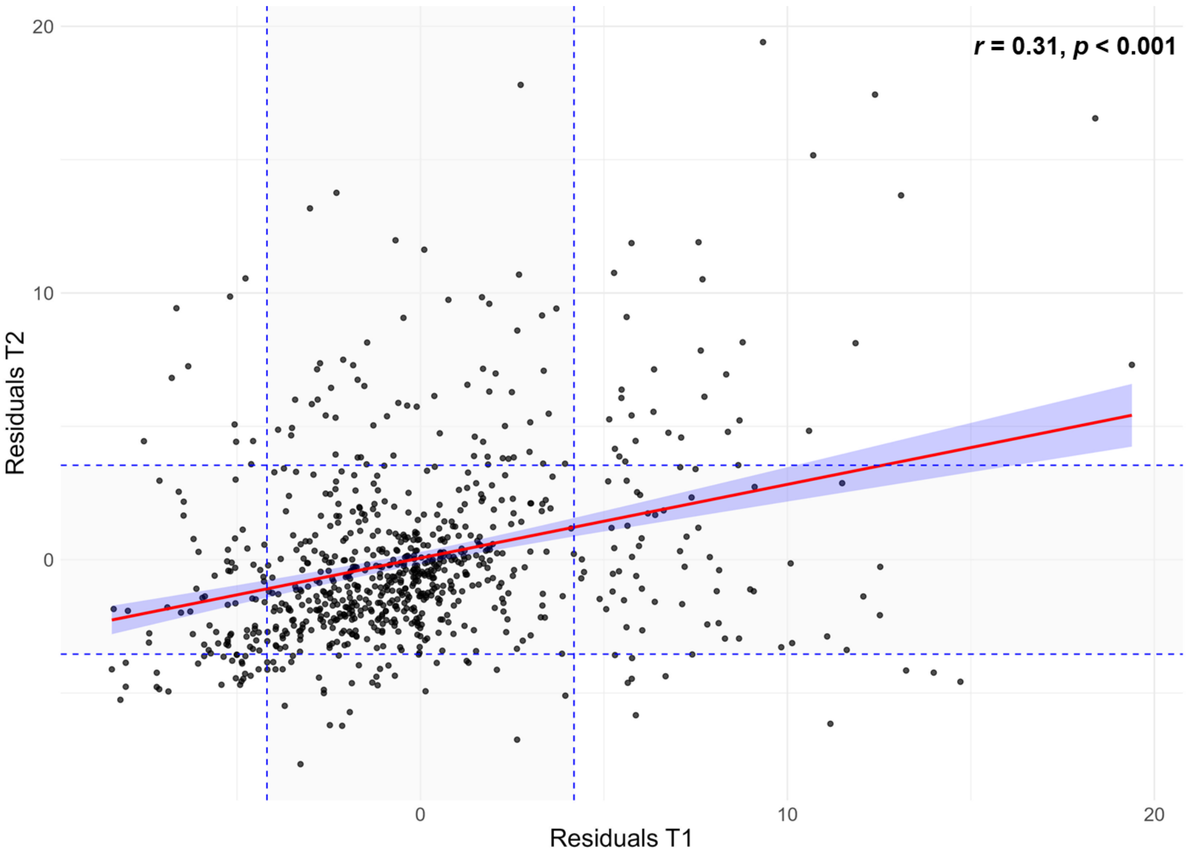


***Note.*** Scatterplot of T1 and T2 residual scores. The regression line (red) shows the significant positive association between time points ($r$ = 0.31, $p$ < 0.001), indicating moderate temporal stability. Dashed lines mark the ±1 residual standard error thresholds used to define resilient and vulnerable extreme groups.

**Supplementary Figure 5.** Impact of Predictor Variables on Outcome at T2.

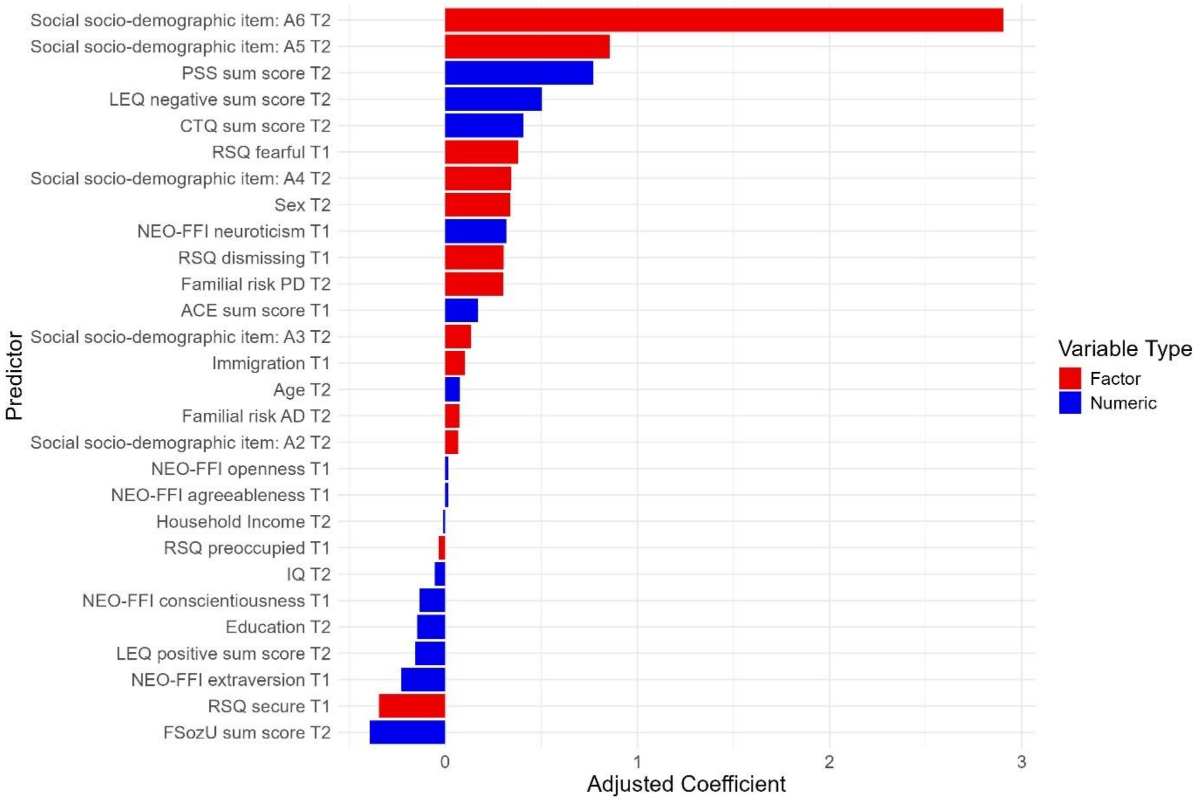


***Note***. Standardized (blue) and unstandardized (red) regression coefficients of all predictors at T2. For continuous predictors, coefficients were scaled by the predictor's standard deviation (SD) and reflect the expected change in HAM-D sum scores per 1-SD increase. Positive coefficients indicate higher predicted depressive symptom severity. Categorical predictors are displayed as unstandardized coefficients representing differences relative to the reference category (e.g., attachment style present vs. absent; familial risk vs. none; female vs. male sex). The ordinal scaled social interaction answer items A2-A6 (increasingly fewer social interactions) reflect differences relative to A1 ("more than one social interaction per week").

**Supplementary Figure 6.** Correlation Matrix of Included Risk and Protective Factors at T2.

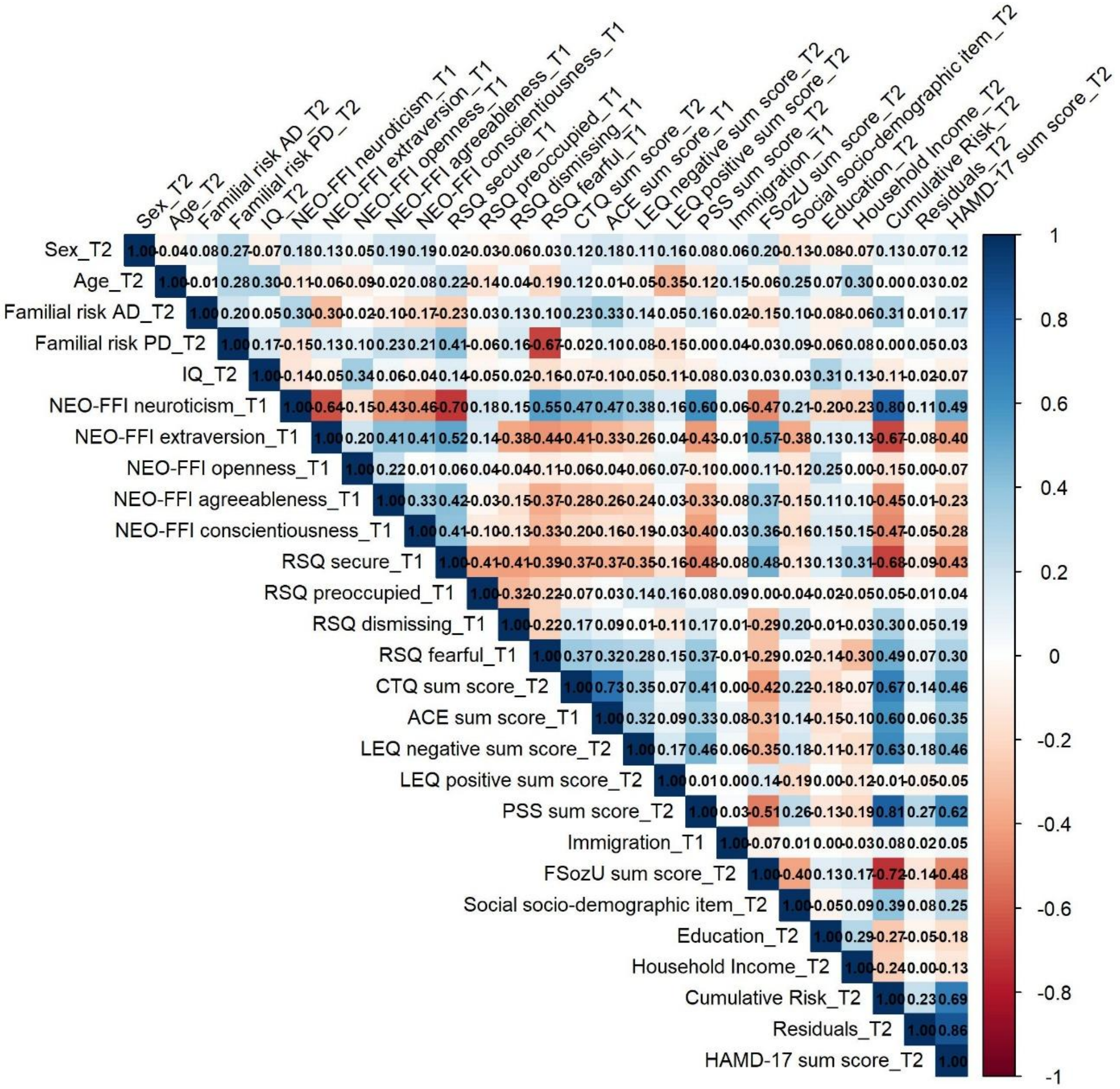


***Note***. Correlation matrix of all predictors at T2, including the 22 risk and protective factors, age, sex, cumulative risk, residual scores, and HAM-D. Pearson correlations were used for continuous variable pairs, polychoric correlations for categorical pairs, and polyserial correlations for continuous–categorical pairs. Values represent correlation coefficients (*r*).

**Supplementary Figure 7.** Resilient Individuals (T1) exhibit Lower GMV at T2 in the left IOFG/Temporal Pole.

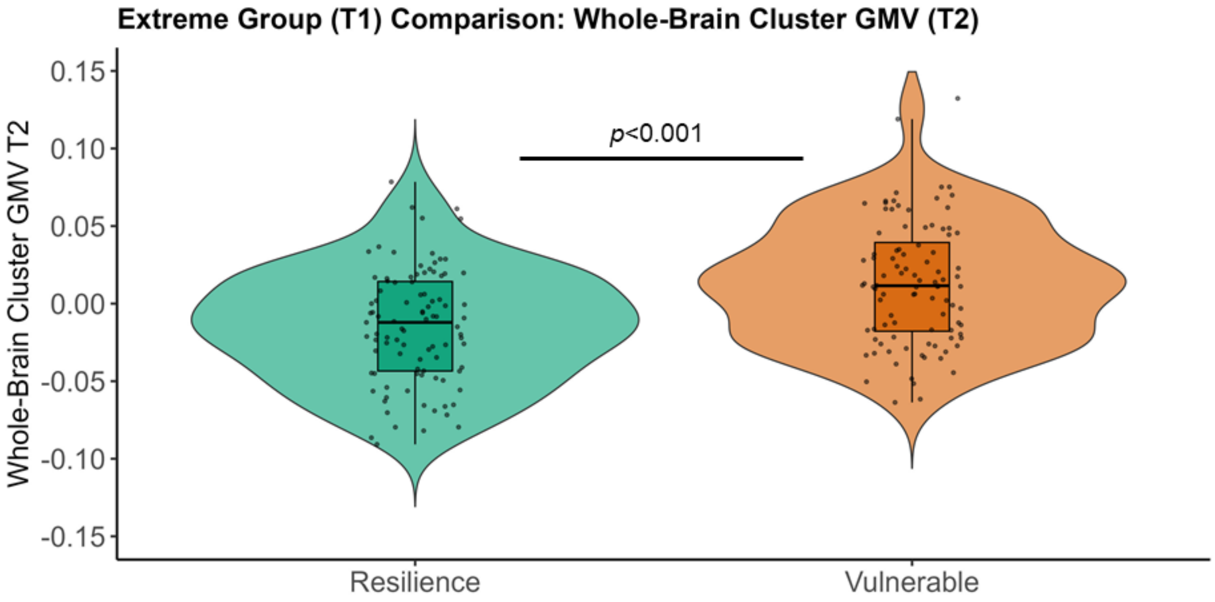


***Note.*** Extreme group comparisons revealed significantly lower gray matter volume (GMV) at T2 (first eigenvariate, partial residuals) in the T1-resilient group (n = 95; green) compared to vulnerable individuals (n = 97; orange) in whole-brain analyses. Whole-brain significant cluster comprises the left inferior orbitofrontal gyrus (IOFG) and left temporal pole ($k$ = 1578 voxels, $p_{FWE(cluster)}$ = 0.007; $d$ = 0.74, peak MNI = −50/18/−12).

**Supplementary Figure 8.** Associations between Resilience-Associated Cluster Gray Matter Volume and Clinical Course Indicators.

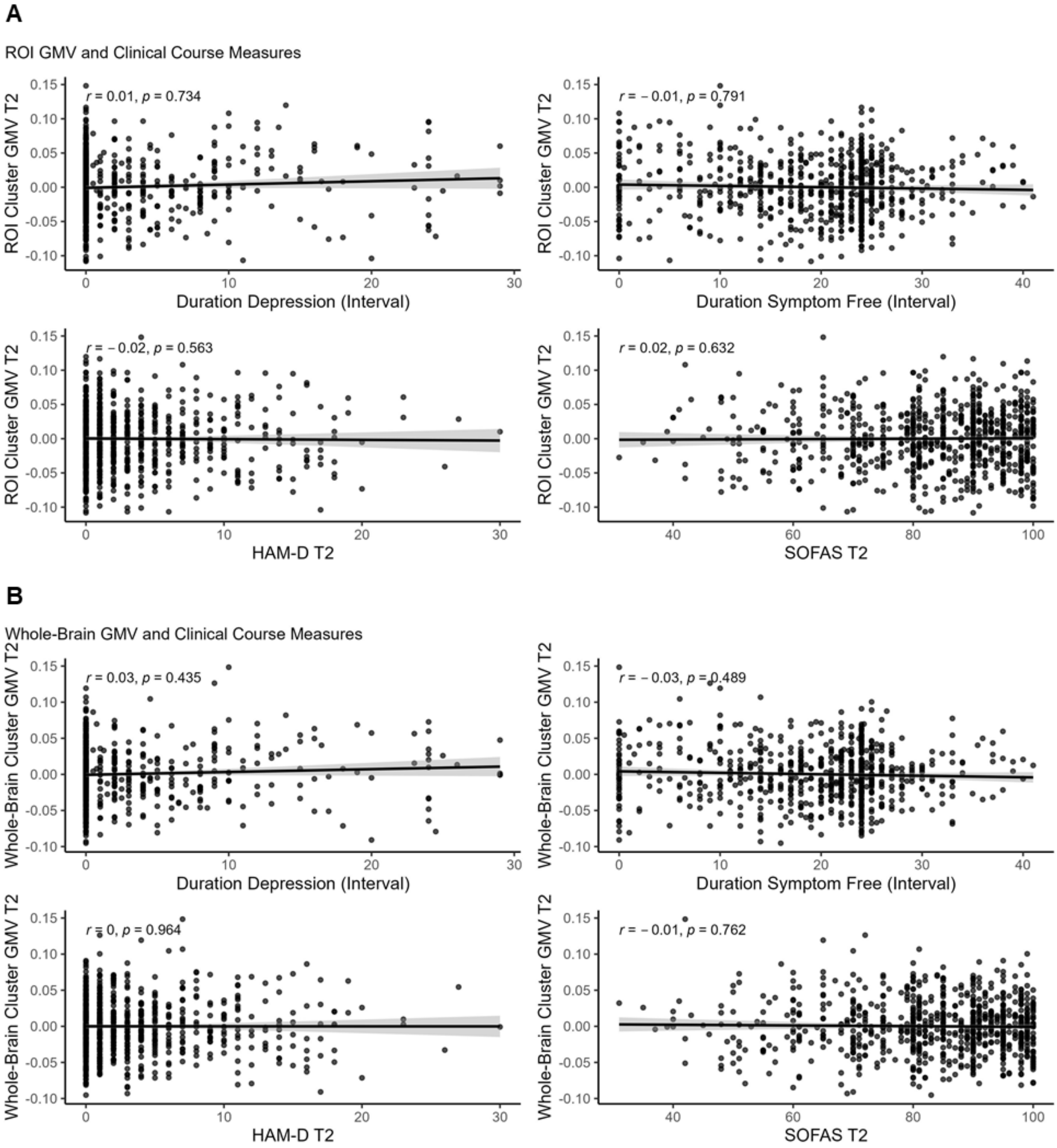


***Note***. Spearman correlations between covariate-adjusted GMV in the resilience-associated left orbitofrontal/temporal-pole cluster at T2 (ROI cluster, A; whole-brain cluster, B) and clinical indicators at T2 (HAM-D, SOFAS) as well as longitudinal course measures (months symptom-free, months depressed). Plots show fitted lines with 95% confidence intervals. No significant associations were observed (all $|r| \leq 0.05$, $p \geq 0.40$).

**Supplementary Material – References**

17. Updegraff JA, Taylor SE (2000): From Vulnerability to Growth: Positive and Negative Effects of Stressful Life Events. *Loss and Trauma*. Routledge.

18. Norbeck JS (1984): Modification of life event questionnaires for use with female respondents. *Res Nurs Health* 7: 61–71.

19. Cristóbal-Narváez P, Haro JM, Koyanagi A (2020): Perceived stress and depression in 45 low- and middle-income countries. *J Affect Disord* 274: 799–805.

20. Keller A, Litzelman K, Wisk LE, Maddox T, Cheng ER, Creswell PD, Witt WP (2012): Does the Perception that Stress Affects Health Matter? The Association with Health

and Mortality. *Health Psychol Off J Div Health Psychol Am Psychol Assoc* 31: 677–684.

21. Cohen S, Kamarck T, Mermelstein R (1983): A global measure of perceived stress. *J Health Soc Behav* 24: 385–396.
22. Lakey B, Orehek E (2011): Relational regulation theory: A new approach to explain the link between perceived social support and mental health. *Psychol Rev* 118: 482–495.
23. Ozbay F, Fitterling H, Charney D, Southwick S (2008): Social support and resilience to stress across the life span: a neurobiologic framework. *Curr Psychiatry Rep* 10: 304–310.
24. Holt-Lunstad J, Smith T, Layton JB (2010): Social Relationships and Mortality Risk: A Meta-analytic Review. *PLoS Med* 7: e1000316.
25. Wang J, Mann F, Lloyd-Evans B, Ma R, Johnson S (2018): Associations between loneliness and perceived social support and outcomes of mental health problems: a systematic review. *BMC Psychiatry* 18: 156.
26. Fydrich T, Geyer M, Hessel A, Sommer G, Brähler E (1999): Fragebogen zur Sozialen Unterstützung (F-SozU): Normierung an einer repräsentativen Stichprobe. [Social Support Questionnaire (F-SozU): Norms of a representative sample.]. *Diagnostica* 45: 212–216.
27. Sommer G, Fydrich T (1991): Entwicklung und Überprüfung eines Fragebogens zur sozialen Unterstützung (F-SOZU). [Development and validation of a questionnaire on social support.]. *Diagnostica* 37: 160–178.
28. McCrae R, Costa P (2008): The five factor model of personality: Theoretical Perspective.
29. Anglim J, Horwood S, Smillie LD, Marrero RJ, Wood JK (2020): Predicting psychological and subjective well-being from personality: A meta-analysis. *Psychol Bull* 146: 279–323.
30. Bucher MA, Suzuki T, Samuel DB (2019): A meta-analytic review of personality traits and their associations with mental health treatment outcomes. *Clin Psychol Rev* 70: 51–63.

37. Costa P, McCrae R (1992): Revised NEO Personality inventory and NEO five-factor inventory (Professional Manual). Odessa: Psychological Assessment Resources. *Psychol Assess Resour Odessa FL*.

43. Steffanowski A, Oppl M, Meyerberg J, Schmidt J, Nübling R (2001): Psychometrische Überprüfung einer deutschsprachigen Version des Relationship Scales Questionaire (RSQ). *Störungsspezifische Ther - Konzepte Ergeb*.

52. Reiss F, Meyrose A-K, Otto C, Lampert T, Klasen F, Ravens-Sieberer U (2019): Socioeconomic status, stressful life situations and mental health problems in children

and adolescents: Results of the German BELLA cohort-study. *PLOS ONE* 14: e0213700.

53. Tang B, Liu X, Liu Y, Xue C, Zhang L (2014): A meta-analysis of risk factors for depression in adults and children after natural disasters. *BMC Public Health* 14: 623.
54. Ridley M, Rao G, Schilbach F, Patel V (2020): Poverty, depression, and anxiety: Causal evidence and mechanisms. *Science*. https://doi.org/10.1126/science.aay0214
55. Sareen J, Afifi TO, McMillan KA, Asmundson GJG (2011): Relationship Between Household Income and Mental Disorders. *ARCH GEN PSYCHIATRY* 68.
56. Thomson RM, Igelström E, Purba AK, Shimonovich M, Thomson H, McCartney G, *et al.* (2022): How do income changes impact on mental health and wellbeing for working-age adults? A systematic review and meta-analysis. *Lancet Public Health* 7: e515–e528.
57. Zaneva M, Guzman-Holst C, Reeves A, Bowes L (2022): The Impact of Monetary Poverty Alleviation Programs on Children's and Adolescents' Mental Health: A Systematic Review and Meta-Analysis Across Low-, Middle-, and High-Income Countries. *J Adolesc Health Off Publ Soc Adolesc Med* 71: 147–156.
58. Andrade N, Ford A, Alvarez C (2021): Discrimination and Latino Health: A Systematic Review of Risk and Resilience. *Hisp Health Care Int*. https://doi.org/10.1177/1540415320921489
59. Masten AS, Lucke CM, Nelson KM, Stallworthy IC (2021): Resilience in Development and Psychopathology: Multisystem Perspectives. *Annu Rev Clin Psychol* 17: 521–549.
60. Bolsinger J, Seifritz E, Kleim B, Manoliu A (2018): Neuroimaging Correlates of Resilience to Traumatic Events—A Comprehensive Review. *Front Psychiatry* 9: 693.
61. Holz NE, Tost H, Meyer-Lindenberg A (2020): Resilience and the brain: a key role for regulatory circuits linked to social stress and support. *Mol Psychiatry* 25: 379–396.
62. Kalisch R, Russo SJ, Müller MB (2024): Neurobiology and systems biology of stress resilience. *Physiol Rev*. https://doi.org/10.1152/physrev.00042.2023
